# The Evolution of Tobacco Marketing to Women and Girls in sub-Saharan Africa

**DOI:** 10.1101/2025.02.27.25323004

**Authors:** Sharon Nyatsanza, Ukoabasi Isip, Omei Bongos-Ikwue, Farida Adamu, Oluwatoyin Christiana Olajide, Adewunmi Emoruwa, Enobong Umoh, Nasara Usman, Ijudaye Shettima

**Affiliations:** National Council Against Smoking, Johannesburg, Gauteng, South Africa; Faculty of Health Sciences, Simon Fraser University, Burnaby, British Columbia, Canada; Gatefield, Abuja, Nigeria; Department of Economics and Social Sciences, Burgundy School of Business, Dijon, Bourgogne-Franche-Comté, France; Centre for Gender and Social Policy Studies, Obafemi Awolowo University, Ife, Osun, Nigeria

## Abstract

**Introduction:** Historically, tobacco use among women and girls in sub-Saharan Africa has been significantly lower than among men. However, recent trends show a concerning rise in smoking rates within this demographic. This shift necessitates a deeper examination of the role tobacco industry marketing plays in driving these changes. Focusing on five key countries—Nigeria, South Africa, Rwanda, Kenya, and Senegal—this research provides a comprehensive analysis of industry marketing tactics targeting women and girls in the region.

**Aims and Methods:** This study aims to investigate the evolving strategies used by the tobacco industry to market products to African women and girls. A mixed-methods approach was employed, combining a literature review, quantitative surveys, and qualitative semi-structured interviews. In addition, a historical analysis of tobacco industry documents and an evaluation of tobacco control laws and regulations in the five surveyed countries were conducted to gain deeper insights into industry practices.

**Results:** Findings from TIDs suggest that the tobacco industry has systematically targeted women for several decades, with a particular focus on young women aged 18-24. None of the surveyed countries currently have comprehensive laws addressing new and emerging products like e-cigarettes. Tobacco marketing was most commonly encountered in nightclubs, bars, lounges, and parties, with 32.8% of participants reporting exposure in these settings. Social media exposure varied across countries, while television shows and movies consistently showed high exposure rates (77.2%) across all five nations. Key informant interviews highlighted dominant themes such as brands targeting females, cultural perceptions of female tobacco use, femininity, autonomy, influencer marketing, digital strategies, harm reduction narratives, proximity marketing, peer and parental influences, and the perceived benefits of tobacco, particularly in terms of flavor, taste, and smell.

**Conclusion and Implications:** The tobacco industry uses sophisticated marketing strategies to enhance product appeal, particularly targeting women through emerging products, flavor manipulation, and harm reduction messaging. Proximity marketing in social settings has proven effective in increasing young women’s access to tobacco products. Critical regulatory gaps remain, particularly concerning e-cigarettes and other novel tobacco products. The adequacy and enforcement of existing TAPS regulations, especially those concerning digital media and cross-border advertising, need urgent attention. Countries should adopt proactive regulations that anticipate industry adaptations and reduce the need for frequent updates. TAPS bans must be extended to encompass emerging tobacco and nicotine products across both traditional and digital platforms. Additionally, regulations need to target proximity and harm-reduction marketing to safeguard young women and prevent the normalization of tobacco use among these vulnerable demographics.

## Introduction

Tobacco use remains a pressing global health crisis, claiming 8 million lives annually, with 80% of deaths occurring in low- and middle-income countries (WHO, 2023). Sub-Saharan Africa (SSA), with its fast growing, youthful population, rising incomes, and expanding tobacco industry (TI) presence, faces a critical public health challenge. Weak regulations, low cigarette prices, and lax enforcement of tobacco control (TC) laws exacerbate this issue (Egbe, 2022).

While historically low, trends reveal that tobacco use among women and girls in SSA is rising, particularly among adolescents, whose smoking rates are nearing those of their male peers (WHO, 2021). This shift calls for an urgent examination of the factors driving these changes, particularly the role of tobacco industry marketing.

Extensive research has been conducted on tobacco marketing to women in high-income countries, including the exploitation of women’s liberation movements in the 1920s (Truth Initiative, 2023). However, little has been done to document similar tactics in SSA. This study aims to contribute to this effort. Investigating the industry’s strategies for targeting women and girls in SSA is crucial for shaping effective policy responses.

By focusing on five key countries—Nigeria, South Africa, Rwanda, Kenya, and Senegal—this research provides a comprehensive, multi-country perspective on industry marketing tactics to women and girls. These countries were chosen due to their significant markets and varying economic contexts within SSA. Additionally, their geographical distribution across different sub-regions of SSA allows for a broad overview of the industry’s marketing activities across various cultural and regional landscapes.

The Framework Convention on Tobacco Control (FCTC) advocates for a comprehensive ban on tobacco advertising, promotion, and sponsorship (TAPS). In February 2024, the Conference of the Parties (COP) to the WHO FCTC adopted specific guidelines addressing cross-border TAPS and the portrayal of tobacco in entertainment media. The COP10 decision emphasized the need for parties to consider restricting or banning TAPS for new and emerging products. The decision also acknowledged the growing role of digital media in transcending national borders and enhancing marketing exposure, particularly among youth, and the need to combat this. Despite being FCTC signatories, the five countries studied still face challenges in regulating tobacco marketing to women and girls, raising questions about industry strategies and tactics.

This study addresses the limited research on female-specific tobacco and nicotine marketing in SSA by analyzing gender-targeted strategies in five countries and explores the marketing of emerging products like e-cigarettes. It incorporates an examination of 30 years of historical internal tobacco industry documents (TIDs), which disclose planned advertising, promotional, and sponsorship strategies within SSA. Furthermore, it investigates the interaction between tobacco marketing tactics and existing TC laws, and the exploitation of digital media to target female smokers, highlighting how these strategies exploit regulatory loopholes. Finally, it provides insights that could guide the development of more robust and effective TC policies.

## Literature review

Understanding tobacco marketing strategies in SSA is essential for crafting effective TC policies. While existing literature largely covers marketing tactics targeting broad demographic groups, there is a notable gap in research on strategies specifically aimed at women. The review examined literature on tobacco marketing tactics across SSA and focused on gender-specific marketing and youth-targeted approaches in SSA.

Although research on female-targeted tobacco marketing is relatively limited, existing evidence suggests that tobacco marketing tactics to women in SSA have been co-opted from those used globally (Goff, 2019). For instance, in South Africa, tobacco advertising often depicts smoking as a symbol of independence and modernity, appealing to young women who aspire to these values (Smith et al., 2018).

Corporate social responsibility (CSR) campaigns comprise a key part of the industry’s strategy to increase its customer base in SSA. For example, in Senegal, Marlboro donated branded school bags to children and sponsored cultural and women’s sporting events, where free samples of tobacco products were given away (Kaleta, 2011). In Kenya, the industry has sponsored community development initiatives aimed at women, and this has subtly promoted tobacco consumption while enhancing its brand reputation (Kinyanjui, 2023).

Harm reduction marketing further plays a key role in targeting women in SSA. In 1998, British American Tobacco began a campaign promoting “light” cigarettes in Southern Africa as part of its regional plan to lure women consumers (Tobacco Tactics, 2020).

Social influencers and covert digital marketing strategies have been used by the industry to bypass laws restricting tobacco advertising. Philip Morris South Africa sponsored a popular female presenter’s trip to Milan Design Week in order to promote IQOS, a tobacco heating device marketed as a healthier alternative to traditional cigarettes to millions in the country (Van-Dyke & Team, 2019). In Kenya, influencers are used to market nicotine pouches to younger audiences (The Guardian, 2021). Tobacco products are often promoted subtly alongside alcohol, food, coffee, and desserts, focusing on visual appeal, which resonates with women’s propensity for such content (Jackler et al., 2020).

In Nigeria, tobacco use is glamorized in films, with female lead characters depicted smoking (Adelufosi, 2014), despite concerns that have led to restrictions of such content (Aguoye, 2024; CAPPA, 2023).

Proximity marketing–locating tobacco sales near areas such as schools and universities–further normalizes tobacco use among youth in SSA, making it accessible and appealing, particularly to girls. The strategic placement of tobacco products near children’s snacks at kiosks and retail stores (Institute for Tobacco Control, 2016; Egbe, 2024) has been noted both in Kenya and Rwanda where e-cigarette kiosks are situated near university campuses (Agaku et al., 2021; Giovenco, 2016) and retail outlets close to schools (Nyirakamana, 2016).

## Methods

### Study Design

This study analyzes the evolution of TI strategies and approaches in marketing to African women and girls over the past 30 years. The marketing analysis examines the industry’s messaging strategy, tactics, and channels targeting African females, including how it leverages cultural, economic, and social trends to reach this growing demographic.

TI marketing strategies across five countries were investigated using a mixed-methods approach, incorporating quantitative surveys of women ranging from age 19 to 75, qualitative semi-structured interviews with female tobacco or nicotine users, as well as teachers, parents, and TC advocates of either gender in these countries.

Qualitative interviews: In selecting participants for qualitative interviews, purposive sampling was used to identify initial respondents. Women who were either consumers of tobacco/nicotine products or exposed to tobacco marketing were identified and recruited. Community leaders, parents/guardians, teachers, and tobacco control advocates were also purposively selected to participate in the interviews, as representing those knowledgeable about social and cultural trends and tobacco use in the community. Snowball sampling was used to identify additional participants. Respondents were drawn from urban and peri-urban areas to capture geographic diversity. Forty-six key informant interviews (KIIs) were conducted. Interview respondents were drawn from each focus country, with women comprising at least 50 percent of interviews.

Quantitative survey/poll: An online survey was administered to a sample of 593 women aged between 19 to 75, from each country, with diverse representation within the age range. Country proportional allocation was used in the distribution of survey respondents, with 153 respondents selected from Nigeria, 120 respondents each from Kenya and South Africa, and 100 each from Rwanda and Senegal. Respondents were identified through social media, email lists and online community forums. Regional stratification was employed, with a focus on urban areas, which have a high media and retail presence. Inclusion criteria was women aged 18-49 who consented to participate. Girls 17 and below were excluded, as well as women who were unable or unwilling to provide written informed consent to participate in the study or to have their data published in ensuing publications.

Ethical approval was sought and obtained from the Federal Capital Territory Health Research Ethics Committee, after submission of the research protocol. The ethics committee also approved the written consent forms used in this study. Written informed consent was obtained for each participant. Consent forms included information about the purpose of the study. Participants were informed that the information collected in the study would be published for use towards potentially strengthening tobacco control policies to promote public health. Participants who agreed to participate were informed that no identifying information would be listed in resulting publications or provided to any third parties, and further gave consent for their statements to be anonymously included in ensuing publications.

The study included a desktop analysis of current tobacco advertising, promotion, and sponsorship (TAPS) policies to evaluate their effectiveness against industry tactics. Additionally, a literature review examined the evolution of marketing strategies, highlighting gender-specific marketing themes and targeted marketing strategies in the selected geographies. The multi-country research team comprises an interdisciplinary group of experts in global health law, tobacco industry marketing, gender, and public health.

### Study Locations

The research focuses on five key countries–Nigeria, Kenya, South Africa, Rwanda and Senegal–selected for their significance as major sub-Saharan African market to the economic interests of the TI, showing an upward trend in tobacco use among target demographic, and their diverse spread across the different sub-regions of SSA. These geographies provide a comprehensive overview of the industry’s marketing activities across diverse geographical and cultural contexts, providing broader insights into the TI’s playbook in SSA.

### Limitations

The study focuses on only five countries within SSA. While these countries represent a diverse set of markets, the findings may not fully capture the state of play in the entire region due to varying economic, cultural, and regulatory contexts. The study is also limited by the lack of weighted country polls which means prevalence results cannot be generalized. Access to comprehensive and current data on tobacco industry marketing strategies over the past 30 years was also limited by small sample sizes and time constraints.

## Results

### Historical Analysis

The TIDs were systematically searched for content related to marketing targeted at women in SSA countries, and the following results were obtained as shown in Table A1.

**Table A1:** Tobacco Documents Keyword Search Criteria.

| S/N | Keywords Search | Number of hits | Number of documents downloaded |
| --- | --- | --- | --- |
| 1 | (((Tobacco) AND (Female)) AND (Marketing) AND (Nigeria)) | 2692 | 10 |
| 2 | (((Tobacco) AND (Female)) AND (Marketing) AND ("South Africa")- title search) | 17 | 3 |
| 3 | (((Tobacco) AND (Female)) AND (Marketing) AND ("Kenya")- title search) | 19 | 0 |
| 4 | (((Tobacco) AND (Female)) AND (Marketing) AND ("Rwanda")- title search) | 0 | 0 |
| 5 | (((Tobacco) AND (Female)) AND (Marketing) AND ("Senegal")- title search) | 1 | 0 |
|  | Total | 2729 | 13 |

Even though the search in Nigeria appeared to have the most documents (10), the South African documents covered the most location-specific details. No country-specific information on female-targeted marketing was identified in documents for Kenya, Rwanda and Senegal. However, there was a significant amount of information on how the TI targets women globally. Some notable themes from the documents were: growth markets and opportunities, brands targeting females, market segmentation by socio-economic status, market segmentation by age-group, harm reduction marketing, cultural perceptions of female tobacco use, and proximity marketing. Table A2 identifies quotes from the TIDs under the major themes. The acronym HORECA represents Hotel, Restaurants and Casinos, while ASU 30 means “Adult Smokers Under 30.”

Findings from the TIDs suggest the TI has systematically targeted women for several decades. In more recent times, women have been considered an important growth market, especially in areas of high population and low smoking prevalence. The most popular age-group of women targeted by the TI in the documents was 18-24 years. Furthermore, the TIDs revealed the industry’s plan to make smoking appear safer for women by introducing low tar and nicotine cigarettes, popularly called “lights.” These “lighter” products are cheaper to produce due to their relatively smaller sizes and thus more profitable when sold in large quantities. The documents revealed targeting of urban women with high socio-economic status with low-tar products to enhance sales in that category. This strategy seemed to be particularly successful in South Africa. In Nigeria, the TIDs identified smokeless tobacco (snuff) as a possible growth market. This suggests the TI was already planning to leverage female preferences for smokeless tobacco in Nigeria and possibly other countries in the region. Furthermore, before comprehensive TAPS bans were introduced, the TI could give the cigarettes a very appealing feminine look. Since TAPS bans are now in place for cigarettes, the TI has shifted their packaging focus to new and emerging products, which, in many SSA countries, have less restrictive or undefined packaging requirements.

### Legal and Policy Analysis

An analysis of the primary tobacco control laws and regulations revealed that none of the countries studied have regulations addressing new and emerging tobacco products. The current definitions of tobacco products are restricted to those containing tobacco leaf, whether wholly or partially, limiting the applicability of laws to non-tobacco products.

Senegal has the strictest tobacco control laws among the surveyed countries, imposing broad bans on both direct and indirect TAPS across platforms. Senegal’s laws prohibit point-of-sale displays, as well as sponsorships and promotional activities. The laws are comprehensive, explicitly banning TAPS across domestic and international television, cross-border channels, internet-based media, and “any medium whatsoever.” This breadth ensures that cross-boundary materials, such as streaming channels and social media, are covered. Senegalese regulations also ban the use, import, advertisement, and sale of shisha or hookah products.

Kenya’s laws prohibit TAPS across various settings including sporting, cultural, artistic, recreational, educational, and entertainment events. However, gaps exist due to the law’s traditional definition of tobacco products and vagueness on CSR activities and sponsorship. The law explicitly prohibits cross-boundary TAPS, banning all communication published or which originates outside Kenya. Kenyan laws also prohibit the sale, use or advertisement of shisha.

South Africa enforces a general ban on TAPS across traditional channels, but allows point-of-sale displays and charitable contributions and sponsorship not aimed at advertising, representing a significant loophole for CSR. The South African law defines promotion to include various means including film, television production, or the internet. A key limitation is that the TAPS ban does not extend to international satellite, film, or video transmission made outside South Africa, leaving room for cross-boundary promotion.

Nigeria has a comprehensive ban on TAPS including point-of-sale displays, vending machines and internet-based communications. Among the countries studied, only Nigeria explicitly bans promotions by media and celebrities, applying directly to influencers. Nigeria’s relatively comprehensive TAPS regulations are weakened by broad exemptions allowing TAPS directed at “consenting persons” aged 18 and over, creating loopholes and ambiguity around marketing in adult-only venues, such as nightclubs and bars. This exemption complicates enforcement of the TAPS ban in settings that should ideally be restricted.

Rwanda’s TAPS regulations are narrow in scope, focusing on traditional restrictions prohibiting free samples, restricting CSR activities by tobacco companies and advertising at cultural and sporting events. However, the Rwanda law does not cover cross-boundary TAPS, point-of-sale advertising or new products, leaving significant gaps. The law also bans the sale, import, use and advertisement of shisha or hookah.

Despite the implementation of some form of TAPS regulations in each country, substantial gaps persist, particularly regarding new and emerging products, which remain unregulated. Additionally, there is a lack of clarity, consistency, and explicitness in laws addressing the evolving media landscape.

### Key Findings from Quantitative Surveys

#### Prevalence and social acceptability of tobacco and nicotine use

A total of 593 female respondents aged 19 to 75, with an average age of 34, were surveyed from the five countries: Nigeria (32.5%), Kenya (16.9%), South Africa (16.9%), Senegal (16.9%), and Rwanda (16.9%). For age distribution, the largest group were those aged 24-28 years (23.6%). Nearly half of the respondents (46.9%) had completed college or university education. The largest group consisted of self-employed women (29.8%), followed by non-government employees and the unemployed, both making up 17.4% of the sample.

The survey examined the prevalence of tobacco and nicotine product use among target demographics across the five countries. Overall, 28.43% of women surveyed have tried or experimented with tobacco and nicotine products (Table 1.0), though prevalence varied widely among the countries, from 0% in Nigeria to 65% in South Africa. Senegal (21%), Kenya (34%), and Rwanda (22%) recorded notable prevalence rates (Table 1.9).

In Nigeria, no respondents reported tobacco use, although other studies indicate low smoking rates among Nigerian women (1.1% in GATS Nigeria 2012). It is important to note that this sample is not weighted, and respondents might have underreported smoking behaviors due to cultural stigma and the social disapproval of smoking in their context.

Cigarettes were the most commonly used tobacco product, with high usage rates reported in Kenya (85%), South Africa (92%), Senegal (78%), and Rwanda (80%). E-cigarettes were notable in South Africa (35%) and Kenya (20%), while shisha use was highest in Senegal (30%) and Kenya (25%). Shisha use is less prevalent in South Africa (18%) and Rwanda (15%) recorded low prevalence of shisha, while Senegal (15%) recorded lower use of e-cigarettes. Nicotine pouches and similar products recorded low usage rates across the board, with the highest prevalence in South Africa (8%) and Kenya (5%), and lowest prevalence in Senegal (3%) and Rwanda (2%) (Table 1.10).

Tobacco use was generally viewed as unacceptable in most of the surveyed countries, with disapproval rates as high as 92% in Senegal (Table 1.17). In Senegal, 92% of respondents view tobacco use as “not at all acceptable,” followed closely by 89% in Rwanda, and 88% in Kenya. In contrast, South Africa showed a significantly different pattern. 35% of respondents in South Africa view tobacco use as “not at all acceptable,” while 5% of respondents consider tobacco use to be “very acceptable,” and 20% view it as “somewhat acceptable.” Besides South Africa, no respondents in the surveyed countries deemed tobacco use “very acceptable” and only 1% saw it as “somewhat acceptable.”

When looking at product usage by age, 29.2% of respondents had tried tobacco or nicotine products. Specifically for e-cigarettes or vapes, the 24-28 age group had the highest usage rate at 15.7%. This was followed by the 29–33-year-olds at 11.2% and the 18–23-year-olds at 9.1%. Usage was lowest among those aged 40 and older, with no reported usage in the 50+ age group. Additionally, e-cigarette or vape usage was highest in South Africa, where 25% of the country’s sample reported usage. College and university graduates had the second-highest usage rate at 12.2%, with post-graduate degree holders reporting the highest rate at 13.6%.

#### Common marketing channels

The survey results provide critical insights into the patterns of tobacco marketing exposure across various channels. Overall, television shows and movies were the dominant marketing channels, with 77.2% of respondents across all five countries encountering tobacco-related content–including advertisements or usage of cigarettes, shisha, or e-cigarettes–through traditional broadcast and streaming platforms such as DSTV (cable TV), Netflix, or Showmax.

Social media marketing was particularly strong in South Africa, where 30% of respondents reported seeing influencers promoting tobacco products. Respondents also encountered product placement in user-generated content (25%), and through giveaways or competitions (18%). Influencer activity was minimal in Kenya with 7% of respondents encountering influencer promotions. The question assessed marketing across various social media platforms, including Facebook, Twitter, Instagram, Snapchat, TikTok, YouTube, and WhatsApp.

Social media exposure to tobacco marketing, including influencers promoting products, giveaways, and product placements, is highest among younger age groups. Individuals aged 18-23 have the highest exposure to online tobacco marketing across all categories: 20.6% were exposed to influencer promotions, 10% to giveaways, and 14.4% to product placements. We found that there was a gradual decline in exposure with age, with respondents above 50 years encountering the lowest exposure (15.8% influencers, 6.6% giveaways, 9.8% product placements). Conversely, 57% of people older than 50 years saw no social media marketing, while the numbers for people aged between 18 and 23 was 51% (Table 1.9).

In-person tobacco marketing was predominantly encountered in nightclubs, bars, and lounges, with 32.8% of participants reporting exposure in these settings. Parties followed closely at 32.2%, while points-of-sale, such as stores and kiosks, accounted for 28.5%. Social events were noted by 22.8% of respondents, and community events showed the least exposure at 8.4%. Additionally, 21.6% of respondents reported no encounters with tobacco marketing. Social events likely refer to more formal or organized gatherings, while parties imply more casual, private settings.

Country-specific insights reveal variations in marketing exposure. In South Africa, the highest exposure occurred in TV shows and streaming channels (84.17%), followed by parties (55.83%), points of sale (50.83%) and social events (46.67%). In Kenya, tobacco marketing was most prevalent on TV shows (84.17%), followed by nightclubs, bars, and lounges (59.17%), parties (42.50%) and points of sale (33.33%).

In Nigeria, TV shows were the primary channel of exposure (84.97%), with lower exposure reported in points of sale (9.80%), nightclubs, bars, and lounges (8.50%) and parties (7.84%). Rwanda followed a similar pattern, with TV shows being the highest exposure source (69%), followed by parties (37%), nightclubs, bars, and lounges (31%), and social events (20%). In Senegal, TV shows also topped the list (64%), with points of sale following closely (34%), and exposure in nightclubs, bars, and lounges (28%) and parties (24%).

### Findings from Key Informant Interviews (Qualitative)

A total of 10 interviews were conducted in Nigeria, 10 in Kenya, 9 in South Africa, 9 in Rwanda, and 7 in Senegal. The following themes and sub-themes were generated to analyze the interviews:

Market segmentation (sub-themes: segmentation by age-group, segmentation by socio-economic status, segmentation by environmental factors); cultural norms (sub-themes - cultural perceptions of female tobacco use, deviant subcultures, Western influence on local cultures, culturally acceptable use of tobacco); users’ perceptions of advertising; Socio-economic status of tobacco users; marketing strategies (sub-themes: Global marketing strategies, regional/local marketing strategies, digital marketing, celebrity or influencer marketing, movies and shows, mainstream media advertisements, point-of-sale ads, event marketing, proximity marketing, billboards, harm reduction marketing, free samples and gifts); users’ perceptions of packaging; growth markets and opportunities; Brands targeting females; feminist ideals and women’s autonomy (sub-themes: agency of choice, women’s autonomy, GBV, social exclusion); Peer and/or parental influence; Pricing of tobacco products; Tobacco regulatory and policy environment; Suggestions for improving tobacco policy; Femininity and acceptability; taste, flavor and smell; perceived harms of tobacco use; perceived benefits of tobacco use.

After extensive analysis of the interview transcripts in Nvivo 14 software, accessed through Simon Fraser University (free trial available at https://lumivero.com/resources/free-trial/nvivo/), A completely free alternative to Nvivo 14 is Qualcoder https://qualcoder.wordpress.com/how-to-use/.

The dominant themes observed across the five countries were:

1. **Brands targeting females:** Historical tobacco industry documents confirm that some brands were made specifically with women in mind as shown in the quote below:

> *“At the other end of the market, the 1985 test launch of Ritz proved very successful. The product appeals particularly to female smokers…Philip Morris is also well positioned with Virginia Slims, the leading ‘female’ brand. National distribution of Virgin Slims 120 at the end of last year helped the brand to achieve a 2.9% market share and exploit the growing relative importance of women to the cigarette industry.”* (ERC Statistics International Ltd., n.d.) Furthermore, the KIIs identified some brands targeting women in SSA such as Velo, a nicotine pouch product in Kenya, Vuse and Modi e-cigarettes (South Africa), and Strawberry cigarettes (Rwanda). Many KII respondents also believed shisha was particularly targeted at women. The KII quote below confirms Strawberry as a female brand for Rwandan women.

> *“Strawberry, most of the time you can see that it attracts a lot of women and the ladies. Because even the design and also how it looks… even the color is attractive for ladies.”* (Rwandan Woman 18-49, 2024)
2. **Cultural perceptions of female tobacco use:** All five study countries have cultures that stigmatize female tobacco use. This led to a lot of female tobacco users being isolated from their families, particularly parents who feel ashamed by their behavior. Many SSA cultures see smoking by women as irresponsible and depicting a lack in moral values:

> *“There is a specific perception regarding women who smoke. People tend to think you are kind of like loose or you don’t have order… This guy sees me smoking - says ‘you have a great personality, I didn’t think you would smoke.’”* (South African Female Smoker, 2024)
3. **Femininity and acceptability:** The use of colors and packaging that appeal to females was a major theme identified across all five countries. Particularly, the use of the color pink, portability of novel tobacco products and the sales of products near other products regularly purchased by women and girls. The quote below corroborates this:

> *“When it comes to packaging, it’s usually the colors that attract people. Certain colors may attract women, or certain aromas…in other cases, you’ll see lip glosses. So, this is specific to attracting women. So, these are strategies to attract … more young women, more young girls.” (Senegalese TC advocate, 2024)*
4. **Women’s autonomy:** To counter the stigma associated with tobacco use, the TI has used feminist ideals to market tobacco as a symbol of choice and freedom:

> *“That the women who smoke are perceived to be tough. They are like men because that’s principally a man’s product and you realize Kenya has had [a] long history of campaigning for the culture - making the culture powerful. So as a way of showing that powerful girl they tend to want to smoke to prove that “we are just like men. We are powerful”* (Kenyan TC Advocate, 2024)
5. **Celebrity/influencer marketing:** The use of celebrities and influencers to market novel tobacco products online and on television has been observed as a strategy to normalize female tobacco use:

> *“Sometimes you find it on Instagram. They would use a famous influencer randomly on Live. She will be smoking a water pipe and probably speaking about a particular flavor… and people start commenting ‘I’d like to try it.’ ‘Where did you get it.’ Or in prominent TV shows they would have a character…that is loved and randomly with [a] vape…and they produce to the population that probably watches that program because of that person who’s on that show. And I think it really does also vary for various groups of girls … or university students that would have parties and probably states that first 20 people to come through will receive a free vape, for all hubbly bubbly (shisha) free for the girls that night at that party.”* (South African Woman 18-49, 2024)
6. **Digital marketing:** With TAPS bans in place in many SSA countries, the TI is exploiting the social media space to promote their products. Tiktok and Instagram were mentioned as possible advertising and promotion channels. Another use of the digital space was for online sales of tobacco products.

Moreover, a KII quote notes that:

> *“The girls in Africa [are] so much into the social media…including advertisements on the social media platforms…and a lot of information …online…has exposed these products to our people.” (Kenyan School teacher, 2024)*

**7) Harm reduction marketing:** Harm reduction marketing has historically targeted women for a number of reasons. Women are more likely to be cautious about health outcomes of their tobacco use than men, who are more likely to exhibit risk-taking behavior. (Uwimana, Okowa and Habtu, 2023; Beia, Kielmann and Diaconu, 2021). Furthermore, increased education among women and girls raises the likelihood that they will opt for tobacco products which appear to be less harmful to their health. Tobacco can adversely affect maternal and reproductive health (Tsiapakidou et al., 2023). The TI thus markets products with lower tar and nicotine as “light” products to women to encourage their use. A TI insider noted the following:

> *“In 1983 52.2% of cigarette smokers opted for medium-tar varieties compared to 44.3% in 1981, whilst the proportion of high-tar consumers fell in the same period from 34.6% to 26.7%. Female regular smokers were more likely than males to smoke cigarettes with a low tar yield; approximately 32% of males smoked cigarettes with 16mg of tar and over compared to 21% of females. The tendency to smoke low tar yield cigarettes is directly related to education, with the better educated preferring lower-tar brands.”* (ERC Statistics International Ltd., n.d.)

The KIIs revealed that many respondents viewed emerging products as being safer than traditional cigarettes. One of the quotes is presented below:

> *“Q: DO you think products like vapes, e-cigs are safer than traditional cigarettes? A: Yes, because they look like softer options of smoking which to my end is good and safe for women and young girls”* (Kenyan Female Tobacco User, 2024)

Historically, the prospects for weight loss have been factors in women’s adoption of smoking (Potter et. al, 2004), reinforced by the marketing of “light” or “slim” cigarettes. Emerging product marketing is picking up this narrative, as highlighted in this KII response:

> *“And it was actually advertising electronic cigarettes where the e-liquid assists you to lose weight. There was also some that had vitamins in them… So, they were introducing this through the e-cigarettes. So, if you miss a meal, you have an e-cigarette that gives a vitamin boost to your body… If you continue to vape these products, it can assist you with losing weight… (South African Female Former Smoker, 2024).*

**8) Movies and TV shows:** To counter the negative image of tobacco use among females, there has been increased depiction of tobacco use in movies and shows across SSA. Some of these are on cable channels like DSTV while others are on streaming sites like Netflix and Showmax. Although tobacco adverts are not shown with these movies, there is a proliferation of smoking scenes in the movies even when they are not essential to the storylines. Nollywood is the world’s third largest film producer, with content widely viewed across the SSA region. A study of 60 Nollywood movies found 73% containing smoking scenes and 78.9% of female lead characters smoking (Adelufosi, 2014). Recent hit movies *Oloture* and *King of Boys,* available on Netflix, show female leads smoking. Such depictions of tobacco use, particularly by women, tends to normalize the public view of smoking as acceptable, and can influence tobacco initiation among women and girls. (Allem et al., 2022) A KII quote corroborates this:

> *“But from South Africa here, there’s a channel [on DSTV] which we call Mzansi. Sometimes…they tell stories based on what is happening locally…and you’ll find that…they will be showing their young girls in beautiful clothes smoking and obviously the young ones which are seeing that… will want to definitely copy…it’s all about and making this cigarette thing as if it’s a cool thing.”* (South African Tobacco Stakeholder, 2024)

**9) Taste, flavor and smell:** As observed earlier, female tobacco use in SSA is highly stigmatized compared to male use. This stigma has led to\ reactions such as many women smoking in secret, away from their families, especially parents. To ensure the secrecy of their tobacco use habits, many female smokers use products that are less detectable and have a more appealing taste, flavor and smell:

> *“Currently, Dunhill is more popular for us and we like to smoke it more than other kinds of cigarettes. They like it because of the smell, no one can know that you smoke. It is a modern one, which does not smell.”* (Rwanda Woman 18-49 VIII, 2024)

> *“Perhaps in perfumes (flavors) you’ll find that some are much more attractive to women than to men. Like strawberry, mint, among others…Cigarettes flavored with menthol, fruit, vanilla or apple are more appealing to women because they like sweetness. The odor is masked, so they don’t smell like cigarettes.”* (Senegalese Female Tobacco User, 2024)

Other notable quotes from the KIIs are provided in table A4.

## Discussion

### Acceptability of tobacco use among women

The quantitative data indicates strong cultural and social resistance to tobacco use among women in most surveyed countries, particularly in Senegal, Rwanda, and Nigeria. In these regions, smoking is heavily stigmatized. However, South Africa, where Western influences are gradually shifting traditional views, emerges as an outlier, showing comparatively higher levels of acceptance of tobacco use. Despite a generally higher acceptance of tobacco in South Africa, females especially among Black and Indian communities, still experience stigma.

In Senegal, female tobacco use is heavily stigmatized, driving women to smoke in private or in nightclubs where they are less likely to be judged. Similarly, in Rwanda, the strong cultural taboo against female smoking often leads to severe social repercussions, including familial rejection and violent harm. In Nigeria, regional differences persist: Northern Nigeria’s adherence to Islamic Sharia law restricts tobacco access, keeping female smoking rates low. In Southern Nigeria, Western cultural influences have led to a greater acceptance of tobacco use among women. These nuanced perspectives reveal a strong cultural resistance to tobacco use in SSA and present an opportunity for the public health community.

Key Informant Interviews show that the TI portrays smoking as a symbol of autonomy and empowerment for women, especially in Kenya, South Africa, and Nigeria. This narrative appeals to women by framing tobacco consumption as a statement of power and independence in male-dominated societies. However, it contains contradictions: tobacco use has traditionally been associated with men, and women’s use of tobacco, as some respondents have submitted, is seen as a challenge to dominant gender socialization. Many women see tobacco use as a way to assert equality with men. Yet, while women’s agency and autonomy of women is a pivotal aspect of gender equality, the industry’s rhetoric manipulates this concept, promoting smoking as a false symbol of empowerment, and misleading women. Men’s risk-taking and poor health-seeking behavior has been attributed to expectations of masculinity which negatively affect life outcomes (Uwimana, Okowa and Habtu, 2023; Beia, Kielmann and Diaconu, 2021). However, this approach undermines long-term health and economic empowerment, exposing women to health risks and financial burdens.

### Regulatory gaps

The legal analysis reveals significant regulatory gaps in addressing new and emerging products. While most countries have bans on conventional cigarettes, e-cigarettes often bypass these restrictions, allowing the tobacco industry to market these products, particularly through digital platforms. In Kenya, Senegal, and Rwanda, laws banning shisha and hookah exist, but regulations on e-cigarettes remain absent. A recurring theme in the KIIs was the significant gap in the regulatory and policy frameworks governing new and emerging products. This aligns with the quantitative findings, which show significant use of e-cigarettes and nicotine pouches among respondents, further normalizing tobacco use. While cigarettes remain the most common tobacco product, e-cigarette use is notably high in South Africa (35%) and Kenya (20%), highlighting a growing acceptance and the need for comprehensive regulations.

The lack of clarity and consistency in laws addressing the evolving media landscape further complicates enforcement. While some countries, like Kenya, ban cross-border TAPS, others, such as South Africa, have exemptions for these materials. The COP 10 guidelines emphasize the need for comprehensive regulations covering all media forms, including films, streaming services, video games, and influencer promotions. As tobacco marketing becomes increasingly digital and global, clear and robust laws will be essential to keep pace with new tactics.

This issue is closely related to the exposure to TAPS in television shows and movies. Survey results reveal that 77.2% of respondents across five countries encountered tobacco-related content—including advertisements and the use of cigarettes, shisha, and e-cigarettes—on platforms like Netflix, DSTV, and Showmax. A significant portion of the content on these platforms is cross-border, indicating a shift beyond traditional broadcast television. Tobacco marketing is now pervasive in streaming services and entertainment media, enabling content to be easily shared across borders, often in areas where such products or marketing practices may be illegal, complicating enforcement efforts.

Updating laws is often burdensome and time-consuming, so countries should adopt forward-thinking regulations. By broadly defining tobacco and nicotine products, including emerging innovations like nicotine analogs, and implementing comprehensive TAPS bans that address evolving digital marketing, countries can minimize the need for frequent revisions. Establishing dedicated teams or consulting experts to monitor industry trends will enable proactive steps. Cooperation between member states and collaboration with international bodies like the WHO FCTC Secretariat can further help anticipate industry changes, resulting in more resilient, long-term regulations.

While having laws in place to regulate cross-border and digital TAPS is crucial, the capacity and availability of financial resources to enforce and regulate digital media has to be assessed. Countries like Kenya may have comprehensive laws banning TAPS on digital platforms, but KIIs reveal stakeholders’ concern about regulators’ capacities to curb tobacco marketing on social media. Merely having regulations is not enough without the technical capacity and/or financial resources to enforce them. This highlights the need for robust regulatory frameworks, enhanced enforcement mechanisms, and adequate technical and financial support.

### Product appeal and targeted marketing strategies

The KIIs demonstrate the interconnected strategies the tobacco industry uses to enhance product appeal, including taste, flavor, smell, harm reduction marketing, and brand targeting. These efforts reflect the tobacco industry’s design to make the use of tobacco and nicotine products more attractive and less stigmatized among women.

In every country studied, the tobacco industry employs brand targeting as a strategic approach to appeal to women by aligning new and emerging products with their preferences and social contexts. Feminine colors, flavors, and packaging—such as pink, purple, and strawberry-flavored products—are used to make tobacco more appealing to female consumers. Some products appear in the shape of lip gloss, nail polish or dolls. Shisha and e-cigarettes, in particular, are marketed as fashionable and discreet, appealing to women in countries like Senegal, where smoking is highly stigmatized. The appeal of these products is enhanced by flavors that mask the unpleasant characteristics of traditional tobacco, making smoking more palatable to women who are sensitive to smell and social perceptions.

The sale of flavored products alongside harm reduction narratives lends increasing acceptance to e-cigarettes. Harm reduction marketing positions products like e-cigarettes as safer, eco-friendly alternatives in a world where people are encouraged to “go green.” KIIs reveal that e-cigarettes are marketed as reducing environmental harm by avoiding cigarette butt waste, though this narrative often misleads consumers about their true environmental impact. Advertisements emphasizing going green are more likely to resonate with women because of socialization. Gender socialization directed to women extols the traditionally feminine values of altruism, compassion and fairness. Women have been shown to care more about climate change than men (Ballew, et. al., 2018). Therefore, when the TI includes messages about “going green”, it uses these factors to its advantage because women assume that by their choice of cigarette, they are contributing to sustainability. This reinforces the need for regulatory oversight of product packaging and beneficial claims.

### Proximity marketing

KIIs highlight the use of proximity marketing, where e-cigarette and tobacco shops are often located near educational institutions, making these products easily accessible to young adults, especially women. This strategy exploits the visibility of these locations to attract younger consumers, a demographic particularly vulnerable to peer pressure and social trends. A South African study revealed that nearly half of surveyed vape shops were located within 5 km radius of educational institutions, correlating with higher e-cigarette use among young adults aged 18– 29 years (Agaku, 2021).

As education gaps close across SSA, with more girls entering higher education, their exposure to tobacco marketing increases. While this implies that advocacy and initiatives towards promoting girls’ education have been effective, the danger of girls and young women’s exposure to tobacco marketing is real. Girls aged 18-24, as the KIIs and tobacco industry documents present, are particularly vulnerable.

According to KIIs, vape shops in South Africa are often strategically located in malls, enhancing product visibility, while in Nigeria, tobacco products are frequently sold in beauty product sections of malls, allowing women to purchase them discreetly, reducing the stigma around buying tobacco. In-person tobacco marketing is most commonly encountered at social venues like nightclubs and bars, where smoking is normalized (Table 1.12). In Kenya, clubs like “1824” cater to younger adults aged 18 to 24 and serve as prime locations for tobacco products promotion, reinforcing the link between smoking and social activities. KIIs reveal that in the African context, women frequent clubs when younger, influencing female targeted marketing strategies. In countries where female tobacco use is stigmatized, women tend to smoke in judgment-free settings like nightclubs, further complicating efforts to reduce tobacco use.

The legal analysis indicates potential loopholes in regulating TAPS in adult-venues. In Kenya and Senegal, where the law prohibits tobacco promotion at various events, including nightclubs and bars, the focus remains on traditional products, leaving gaps for the marketing of new and emerging products. Nigeria’s TAPS regulations include a broad exemption for “consenting persons” aged 18 and over, further complicating enforcement in adult-only venues like nightclubs and bars. South Africa permits point-of-sale displays and vending machines in adult-only venues, creating another regulatory gap. To address these gaps, laws need to cover new and emerging products, and regulations on sponsorship and promotions need to be comprehensive. They should encompass all forms of corporate social responsibility (CSR) and advertising across various events, including those held at adult-only venues.

Additionally, the existence of designated smoking sections in venues like bars and nightclubs across the surveyed countries raises concerns about the practicality of enforcing comprehensive TAPS bans in areas that are not 100% smoke-free. This calls for further research into effective enforcement strategies.

### Digital, celebrity/influencer marketing

KIIs show that digital platforms and female influencers play a significant role in promoting tobacco products, particularly shisha, e-cigarettes, and other emerging products. Social media campaigns often feature influencers endorsing these products, positioning them as trendy and desirable. A KII insight reflects this approach: “I feel like the poster child of shisha [were] the female influencers” (Kenyan Female Tobacco User, 2024). Another informant in Rwanda observed that “Facebook, Instagram, TikTok” play a role in increasing tobacco use among youths. (Rwanda Woman 18-49 V, 2024).

In South Africa, KIIs confirm the prevalence of influencer marketing, with platforms like Instagram and TikTok being used to promote new products. In Nigeria, stakeholders highlighted the changing media landscape: “…*Not many people would notice that tobacco products are not advertised on billboards or on TV, because we really do not look at billboards or TV, compared to social media. If you spend one hour watching TV or looking at billboards, you would spend four hours on social media being inundated by advertising content. So, that social media loophole is a big minus for TAPS…”* This insight is corroborated by quantitative data, which shows significant levels of digital marketing engagement.

South Africa, Senegal, and Kenya have explicit bans on social media marketing for traditional tobacco products, but these regulations do not adequately address the nuances of digital marketing for emerging products such as e-cigarettes. Nigeria’s regulatory framework, which exempts tobacco advertising aimed at consenting adults, complicates enforcement and allows potential online marketing. Rwanda’s focus on shisha advertising bans, leave gaps in regulating digital marketing for other tobacco products.

Quantitative data shows varied exposure to social media marketing: 18.13% of respondents saw influencers promoting tobacco products, 12% encountered product placements, and 8.23% were exposed to giveaways or competitions. Yet, over half of respondents (54.13%) reported no exposure to tobacco marketing on social media, reflecting varying levels of marketing reach (Table 1.4). Both KIIs and quantitative data underscore widespread digital media and influencer marketing of tobacco products. These figures highlight the growing of digital platforms in tobacco advertising, calling for updated and more comprehensive regulations across SSA.

## Summary and conclusion

### Marketing Strategies

The tobacco industry employs highly sophisticated marketing strategies to target women in sub-Saharan Africa, leveraging cultural, social, and economic trends to make tobacco and nicotine products more appealing. These strategies include flavor and taste manipulation, harm reduction narratives, and proximity marketing in places frequented by young women. Emerging products, such as e-cigarettes and nicotine pouches, are marketed as safer alternatives and position particularly new and emerging products as female friendly.

### Regulatory Gaps

Significant regulatory gaps exist, particularly regarding the control of new and emerging products like e-cigarettes. While traditional tobacco products face varying levels of regulation, emerging products often escape restrictions, allowing the industry to target vulnerable populations, including women and youth, through digital platforms and social media. Cross-border and digital TAPS remain inadequately regulated in many countries, allowing influencers and online content to circumvent existing laws. Furthermore, the impact of weakened regulatory departments and agencies due to corruption, lack of funding and political will, on women’s tobacco use is evident with the sale of illicit tobacco products and sales of tobacco sticks to women.

### Cultural Resistance and Shifting Norms

The research reveals significant cultural resistance to tobacco use among women across Sub-Saharan Africa (SSA), in countries like Senegal, Rwanda, and Nigeria (particularly Northern Nigeria). However, norms are gradually shifting in places like South Africa and Southern Nigeria, where Western influences and tobacco industry messaging present smoking as a symbol of empowerment. The positioning of tobacco as a symbol of empowerment and autonomy risks undermining the unacceptability of tobacco use.

### Prevalence

Prevalence data reveals significant regional variations in tobacco use. South Africa shows higher tobacco use rates, with substantial use of both traditional and emerging products like e-cigarettes. In contrast, countries like Senegal and Rwanda report lower levels of tobacco use among women, reflecting strong cultural resistance. However, the growing acceptance of new and emerging tobacco products highlights the need for updated regulatory measures.

### Proximity Marketing

Proximity marketing, with tobacco products placed near schools, universities, and popular social venues, has effectively normalized tobacco use among young women in several countries. Digital marketing, particularly through influencers and social media, also plays a critical role, making tobacco products visible and desirable to a younger audience. This presents a significant challenge for regulators, as digital platforms continue to be inadequately controlled in many SSA countries.

Findings from this research highlight the pervasive strategies employed by the tobacco industry to target women across SSA. This trend is facilitated by significant regulatory gaps, particularly around digital media, cross-border TAPS and some new and emerging products. Despite strong cultural resistance in many SSA countries, changing norms in places like South Africa pose new challenges. The challenge now is to address the marketing tactics that erode these cultural resistances and prevent the normalization of tobacco and nicotine use among women. The use of CSR by the tobacco industry further raises the issue of gaps which exist in the economic context. Women in particular are confronted with multiple layers of inequality from the private to the public sphere (African Development Bank, 2020). The concentration of TI-sponsored CSR projects in rural areas where awareness of tobacco health harms are lacking presents a threat of increased tobacco use in those areas.

## Recommendations

1. **Adopt Gender-Responsive Approaches:** Tobacco control policies should account for the unique ways the industry targets women and girls. These gender-sensitive policies must go beyond general regulations to include strategies that address gender-specific marketing, such as a restriction on marketing flavored products in colorful packaging and proximity marketing near schools and universities. Regulations should address the unique vulnerabilities of this demographic, particularly through information, education, and communications campaigns focused on empowering women to make informed health choices. Because new and emerging tobacco products are intentionally designed to appeal to feminine sensibilities, policymakers must introduce regulations mandating the introduction of labels showcasing their harmful effects to curb harm-reduction communication tactics.
2. **Strengthen and Adopt Proactive Legal Frameworks:** Countries should revise and update laws and regulations to restrict the promotion of new and emerging products. TAPS bans should be extended to address the aggressive marketing of these products, particularly on social media and other digital platforms where traditional tobacco advertising restrictions fall short. This includes closing loopholes that allow tobacco companies to exploit unregulated spaces, such as influencer promotions and digital giveaways, which are rapidly normalizing these products among women and youth. Countries should also adopt regulations that anticipate industry adaptations and reduce the need for frequent updates. Cooperation among member states and collaboration with international bodies like the WHO FCTC Secretariat will enhance the ability to anticipate changes, resulting in more resilient, long-term regulations.
3. **Strengthen Budget Advocacy for TC Resources:** Tobacco control programs are often under-resourced. Funding is necessary to ensure the strong enforcement of policies, strengthen awareness creation, and enable multi-sectoral collaboration. Lean tobacco control budgets hinder these operations and embolden tobacco companies to introduce CSR initiatives that provide platforms to promote their products and undermine public health efforts. By intensifying budget advocacy efforts, civil society and other tobacco control advocates can help to secure local funding for tobacco control.
4. **Strengthen Cross-Border and Digital Advertising Controls:** Tobacco marketing no longer adheres to national boundaries, especially with the rise of digital media. Countries across SSA should collaborate to regulate cross-border TAPS, creating a unified front against tobacco advertising that crosses national borders through digital platforms, streaming services, and influencer-driven content. Regulatory bodies should exchange information and coordinate enforcement efforts to ensure that tobacco companies cannot exploit weak points in the system. Countries should regularly update and evaluate their tobacco advertising laws to keep up with changes in media and marketing in line with the COP 10 Specific guidelines on Article 13. This includes stricter controls on tobacco depictions in movies, social media, and video games, with special attention to rural and underserved areas where awareness of tobacco harms is limited.
5. **Counter the Industry’s Empowerment Narrative**: Public health campaigns must reframe the industry’s false narratives of empowerment to mean prioritizing health and real freedom. Feminist advocacy must promote ideals that de-center men and the wide range of risky masculinist behavior and performance. This includes messaging that challenges harmful narratives of autonomy tied to smoking. Campaigns should emphasize that tobacco use, far from being a symbol of freedom, exposes women to financial strain and health risks. This narrative shift is crucial to breaking the cultural association between smoking and empowerment, especially in contexts where the TI targets women under the guise of liberation. A behavioral change approach that frames men’s risk-taking behavior as a detrimental pattern can serve to reduce smoking prevalence among men; this can produce a domino effect of curbing smoking among women. With a narrative that downplays smoking as a masculine indulgence, women are less likely to perceive smoking as aspirational. Alongside this, women’s health-seeking inclinations and the feminine ideal of self-care should be emphasized

## Data availability statement

https://doi.org/10.5281/zenodo.14028939

## Annex

### Survey Results

Overall summaries for key data points without age and country segmentation

**Table 1.0.**
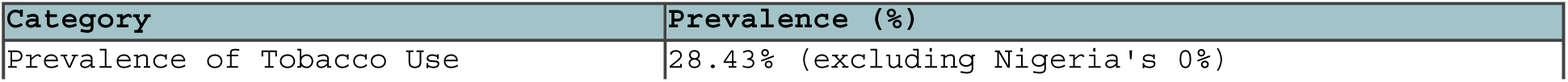
Prevalence of Tobacco Use.

| Category | Prevalence (%) |
| --- | --- |
| Prevalence of Tobacco Use | 28.43% (excluding Nigeria's 0%) |

**Table 1.1.**
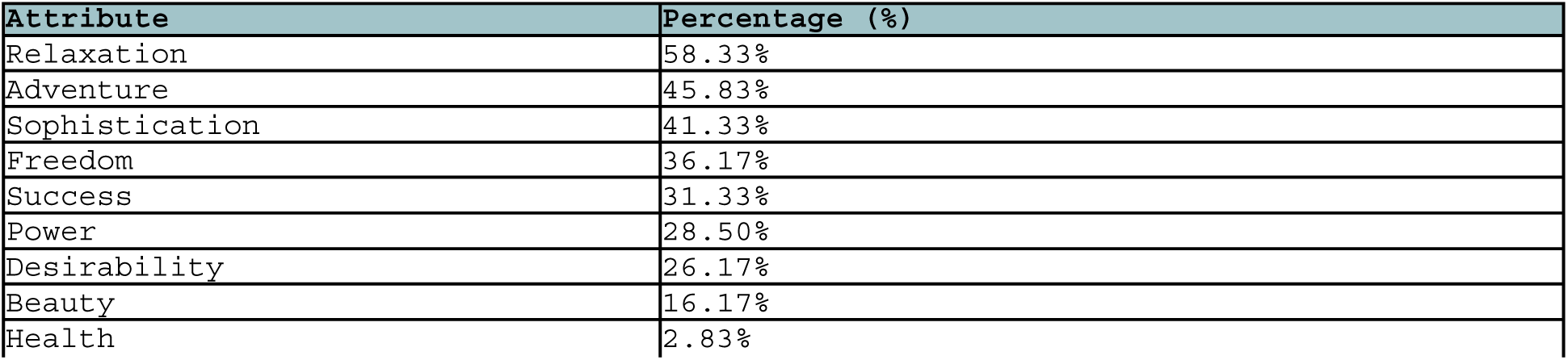
Perception of Tobacco Advertising Attributes.

| Attribute | Percentage (%) |
| --- | --- |
| Relaxation | 58.33% |
| Adventure | 45.83% |
| Sophistication | 41.33% |
| Freedom | 36.17% |
| Success | 31.33% |
| Power | 28.50% |
| Desirability | 26.17% |
| Beauty | 16.17% |
| Health | 2.83% |

**Table 1.2.**
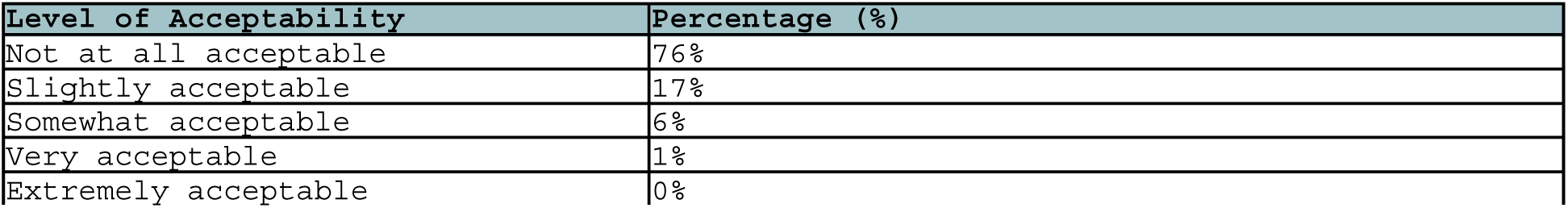
Acceptability of Women Smoking.

| Level of Acceptability | Percentage (%) |
| --- | --- |
| Not at all acceptable | 76% |
| Slightly acceptable | 17% |
| Somewhat acceptable | 6% |
| Very acceptable | 1% |
| Extremely acceptable | 0% |

**Table 1.3.** Tobacco Use by Education and Employment.

| Category | Prevalence (%) |
| --- | --- |
| Self-employed | 38% |
| Employed | 33.5% |
| Unemployed | 25% |
| Students | 20% |

**Table 1.4.**
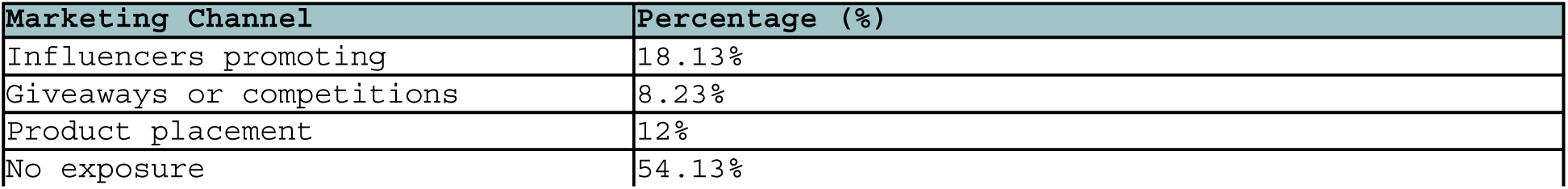
Social Media Marketing Exposure.

| Marketing Channel | Percentage (%) |
| --- | --- |
| Influencers promoting | 18.13% |
| Giveaways or competitions | 8.23% |
| Product placement | 12% |
| No exposure | 54.13% |

**Table 1.5.** Marketing Channels Exposure.

| Channel | Percentage (%) |
| --- | --- |
| Nightclubs, Bars, Lounges | 35.3% |
| Parties | 33.37% |
| Points of Sale | 31.27% |

**Table 1.6.** Exposure Through TV Shows and Movies.

| Channel | Percentage (%) |
| --- | --- |
| TV Shows, Movies, Streaming | 77.2% |

**Table 1.7.** Overall Exposure Through Social Media.

| Channel | Percentage (%) |
| --- | --- |
| Overall Social Media Exposure | 45.87% |

**Table 1.8.** Prevalence of Tobacco Use by Education Level.

| Education Level | Prevalence (%) |
| --- | --- |
| No formal schooling/Primary school | ~15% |
| Secondary school completed | ~30% |
| College/university completed | ~40% |
| Post-graduate degree | ~35% |

**Table 1.9.** Prevalence of Tobacco and Nicotine Use.

| Country | Kenya | SA | Senegal | Rwanda | Nigeria |
| --- | --- | --- | --- | --- | --- |
| Prevalence of Tobacco Use (%) | 34.2% | 65% | 21% | 22% | 0% |

**Table 1.10.** Product usage among those who have tried tobacco products.

| Product | Kenya | SA | Senegal | Rwanda | Nigeria |
| --- | --- | --- | --- | --- | --- |
| Cigarette | 85% | 92% | 78% | 80% | 0% |
| E-cigarettes or Vapes | 20% | 35% | 15% | 0% | 0% |
| Shisha | 25% | 18% | 30% | 15% | 0% |
| Nicotine Pouch | 5% | 8% | 3% | 2% | 0% |
| Nicotine/Lozenge/Gummies | 2% | 5% | 1% | 1% | 1% |
| Others | 3% | 2% | 1% | 1% | 0% |

**Table 1.12.** Marketing Exposure Channels.

| Country | Nightclubs, Bars, Lounges (%) | Parties (%) | Points of Sale (%) | No Exposure (%) |
| --- | --- | --- | --- | --- |
| South Africa | 50.8% | 55.8% | 50.8% | 15% |
| Senegal | 28% | 24% | 34% | 36% |
| Rwanda | 31% | 37% | 31% | 31% |
| Nigeria | 8.5% | 7.8% | 9.8% | 15% |
| Kenya | 59.2% | 42.5% | 33.3% | 15.8% |

**Table 1.13.** Encounter with Tobacco Adverts on TV and Streaming Platforms like Showmax and Netflix.

| Country | Kenya | SA | Senegal | Rwanda | Nigeria |
| --- | --- | --- | --- | --- | --- |
| Yes | 84.17% | 84.17% | 64% | 69% | 84.97% |
| No | 15.83% | 15.83% | 36% | 31% | 15.03% |

**Table 1.14.** Social Media Marketing Exposure.

| Country | Influencers Promoting Tobacco/Vapes/Nicotine/Shisha | Giveaways or Other Competitions | Product Placement in User-Generated Content | No Exposure (%) |
| --- | --- | --- | --- | --- |
| South Africa | 30% | 18% | 25% | 27% |
| Senegal | 7% | 3% | 5% | 85% |
| Rwanda | 10% | 5% | 8% | 77% |
| Nigeria | 8.50% | 7.84% | 9.80% | 15.03% |
| Kenya | 15% | 8% | 12% | 65% |

**Table 1.15.**
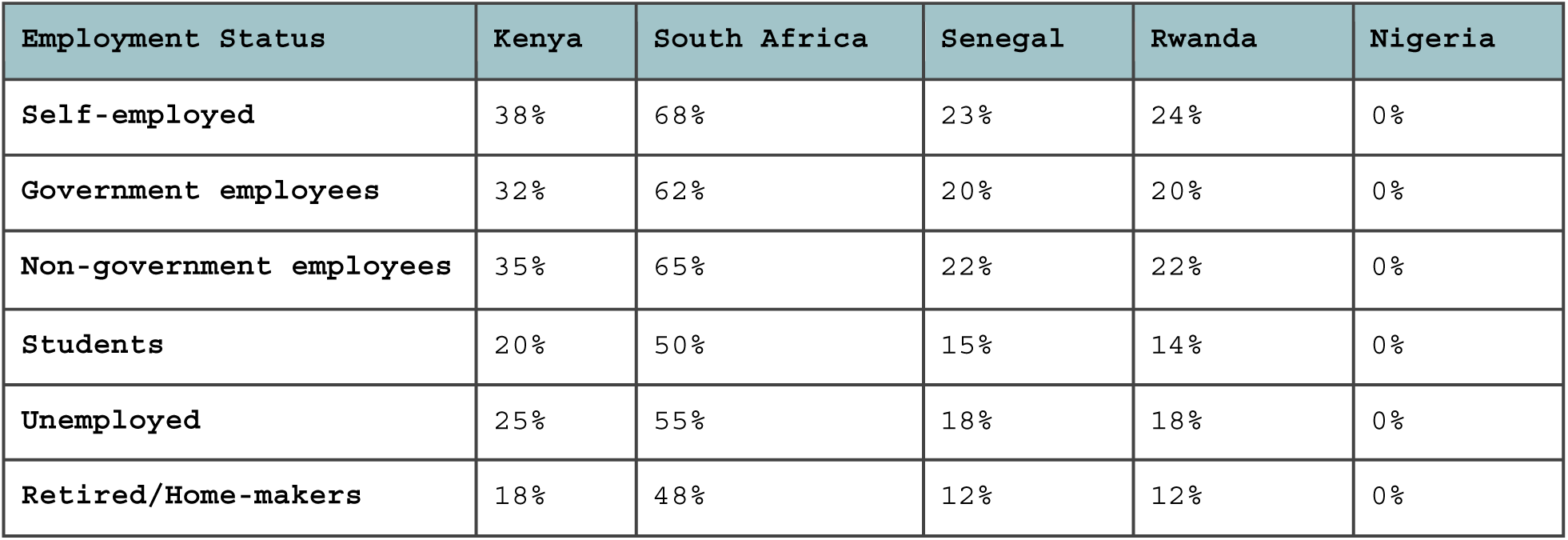
Tobacco Use Prevalence by Employment Status.

**Table 1.16.** Tobacco Use Prevalence by level of Education.

| Education Level | Kenya (%) | South Africa (%) | Senegal (%) | Rwanda (%) | Nigeria (%) |
| --- | --- | --- | --- | --- | --- |
| No formal schooling/Primary school | 15% | 45% | 10% | 10% | 0% |
| Secondary school completed | 30% | 60% | 20% | 18% | 0% |
| College/university completed | 40% | 70% | 25% | 25% | 0% |
| Post-graduate degree | 35% | 65% | 22% | 22% | 0% |

**Table 1.17.** Perceptions of acceptability for Tobacco use by country.

| Acceptability Level | Kenya (%) | South Africa (%) | Senegal (%) | Rwanda (%) | Nigeria |
| --- | --- | --- | --- | --- | --- |
| Not at all acceptable | 88% | 35% | 92% | 89% | 0% |
| Slightly acceptable | 11% | 40% | 7% | 10% | 0% |
| Somewhat acceptable | 1% | 20% | 1% | 1% | 0% |
| Very acceptable | 0% | 5% | 0% | 0% | 0% |
| Extremely acceptable | 0% | 0% | 0% | 0% | 0% |

**Table 1.18.**
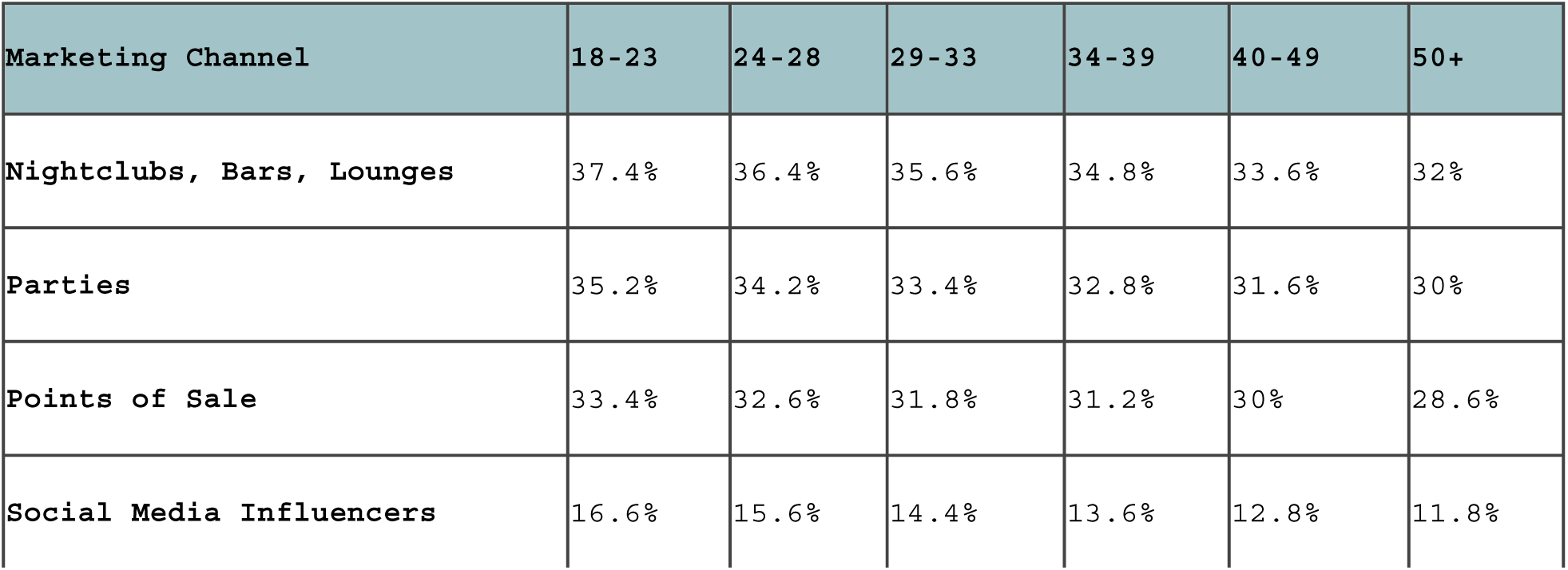
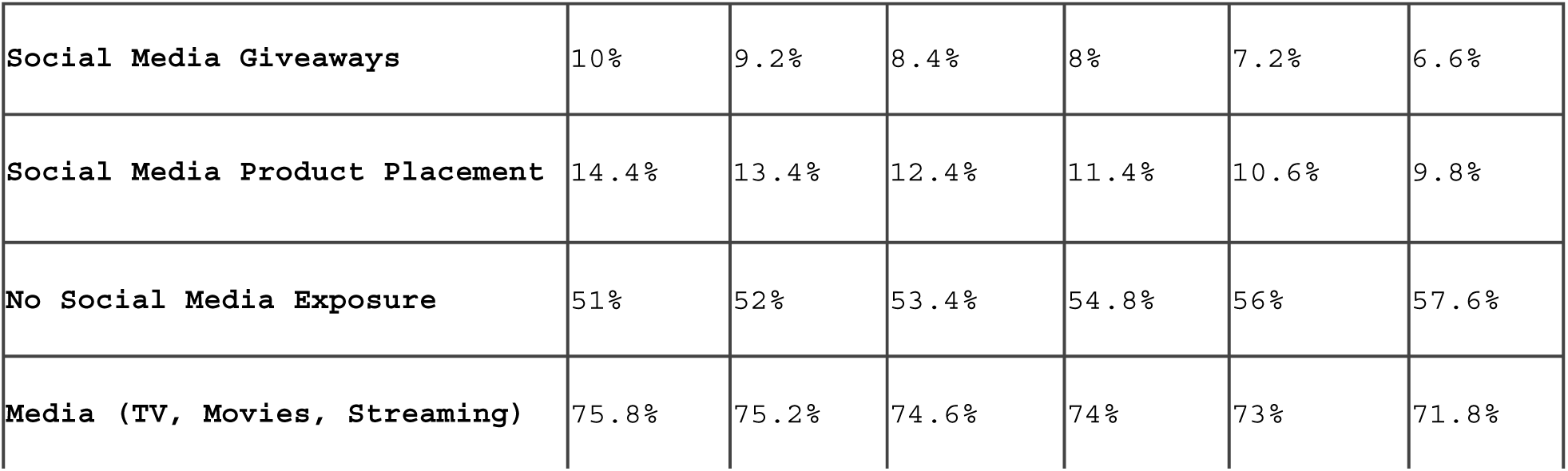
Marketing Exposure by Age Group.

**Table 1.19.**
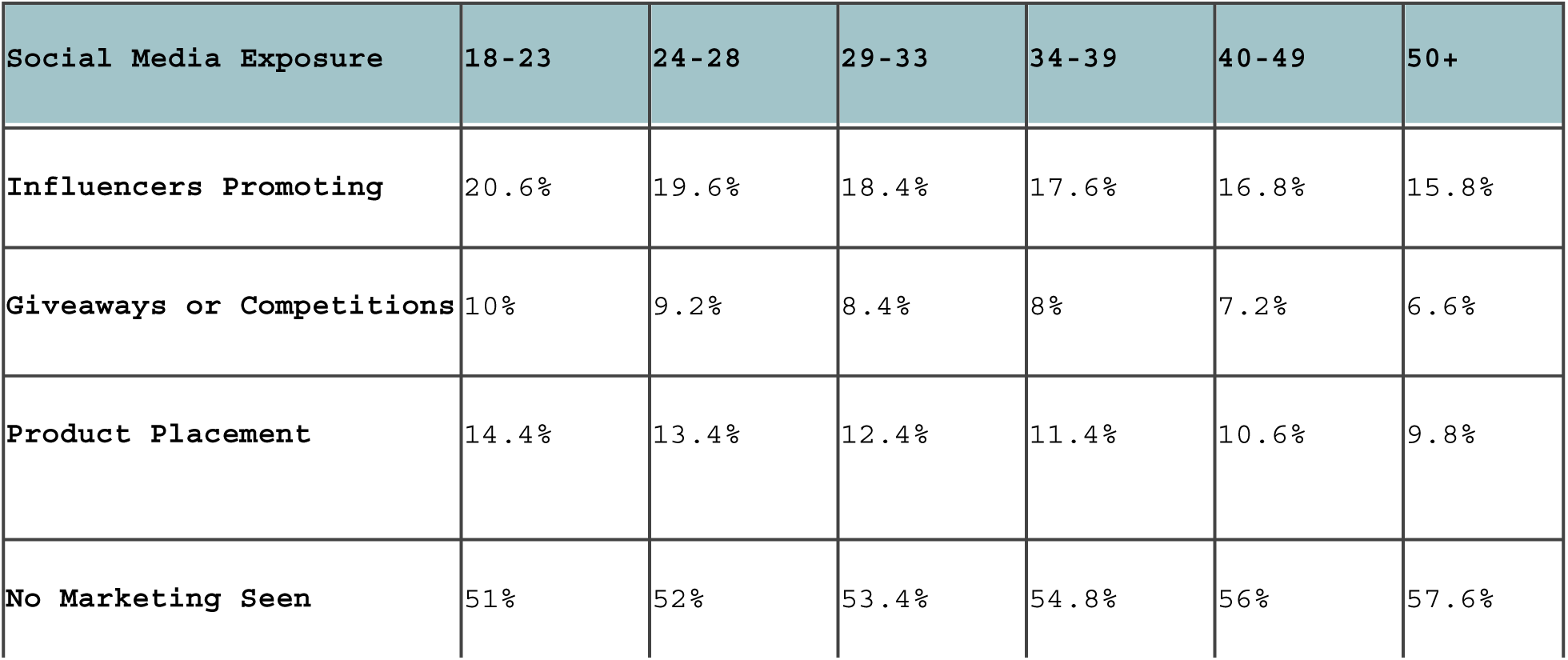
Social Media Exposure by Age Group.

| Social Media Exposure | 18-23 | 24-28 | 29-33 | 34-39 | 40-49 | 50+ |
| --- | --- | --- | --- | --- | --- | --- |
| Influencers Promoting | 20.6% | 19.6% | 18.4% | 17.6% | 16.8% | 15.8% |
| Giveaways or Competitions | 10% | 9.2% | 8.4% | 8% | 7.2% | 6.6% |
| Product Placement | 14.4% | 13.4% | 12.4% | 11.4% | 10.6% | 9.8% |
| No Marketing Seen | 51% | 52% | 53.4% | 54.8% | 56% | 57.6% |

**Table A2:**
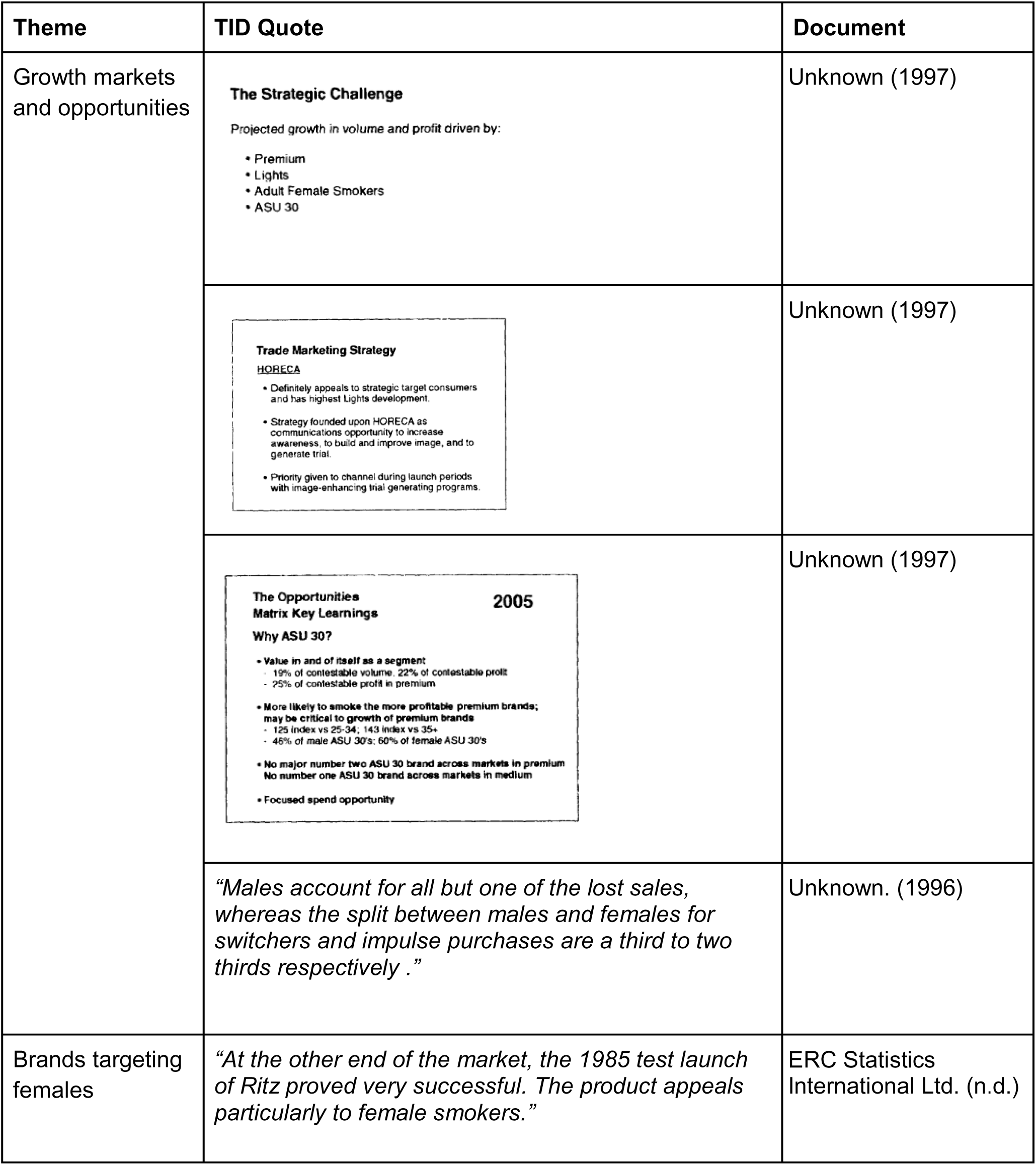
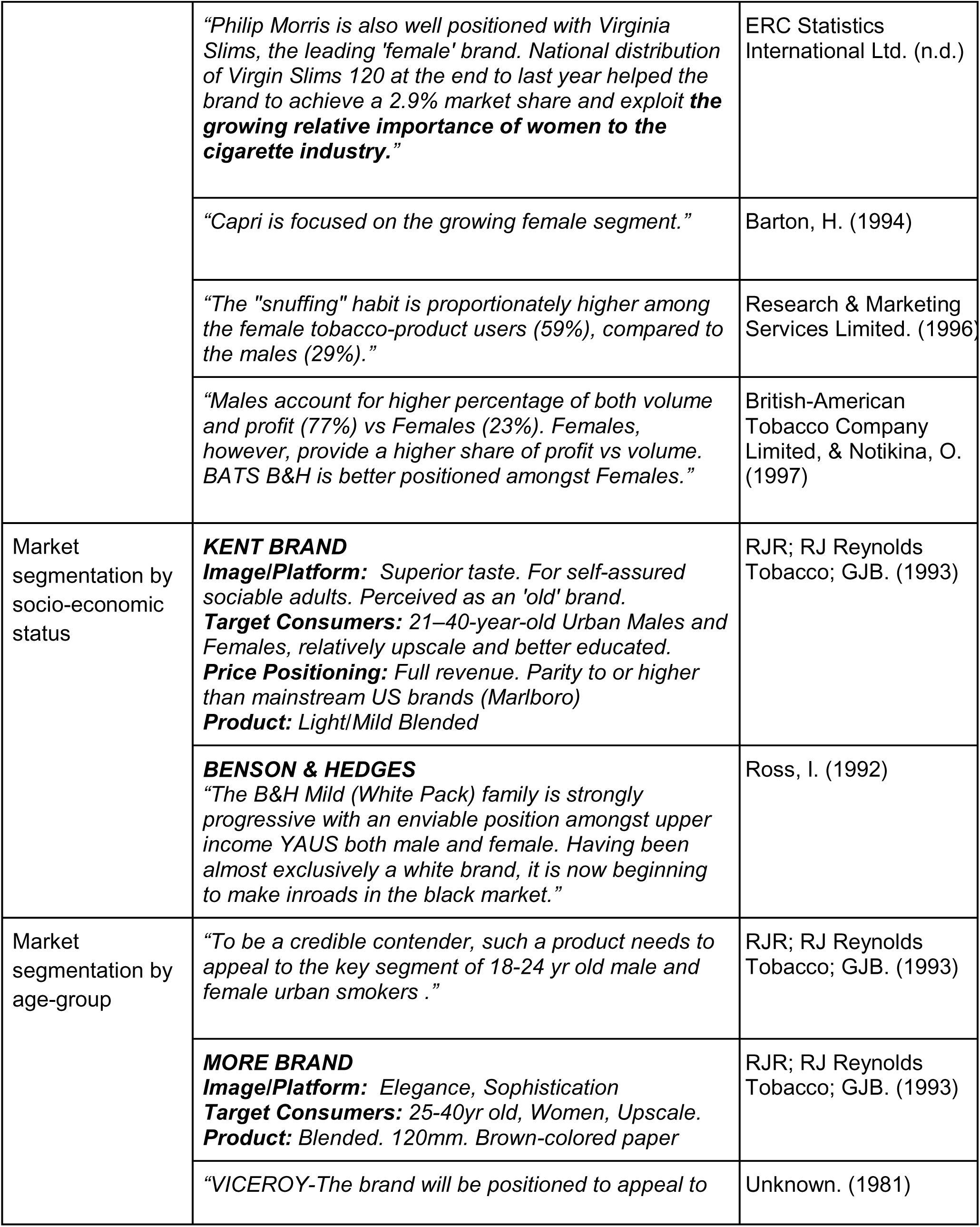
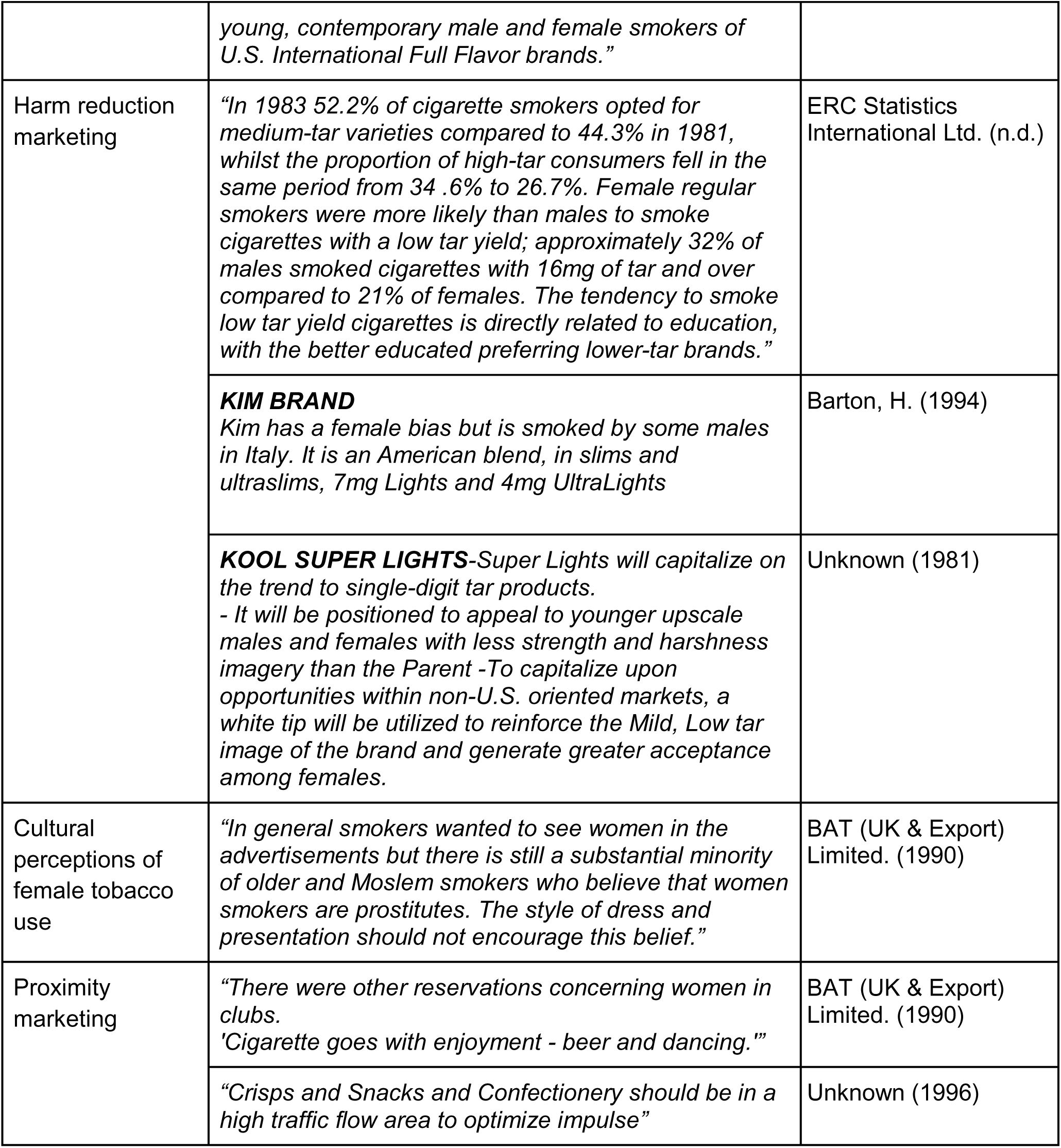
Results of TID search.

**Table A4:**
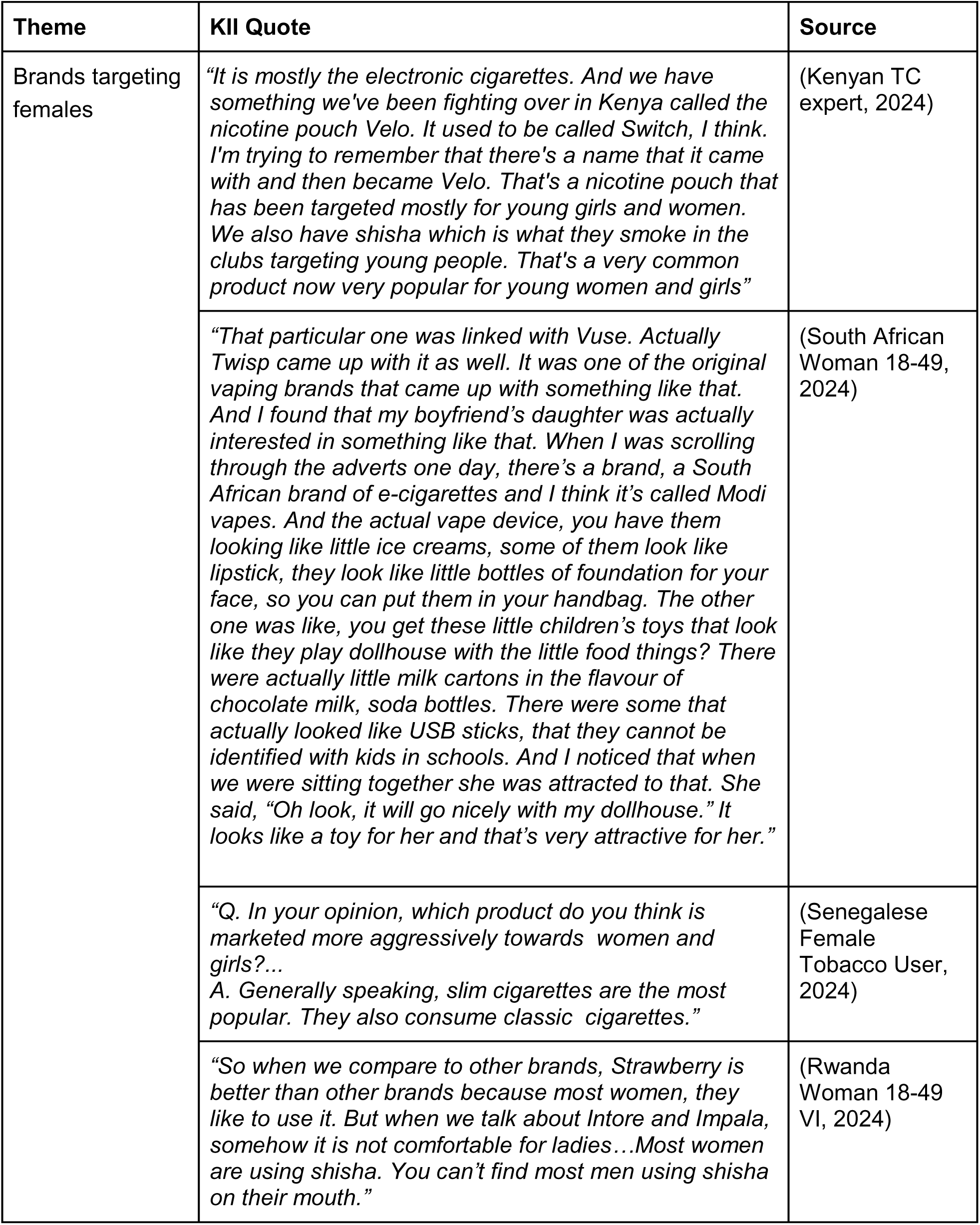
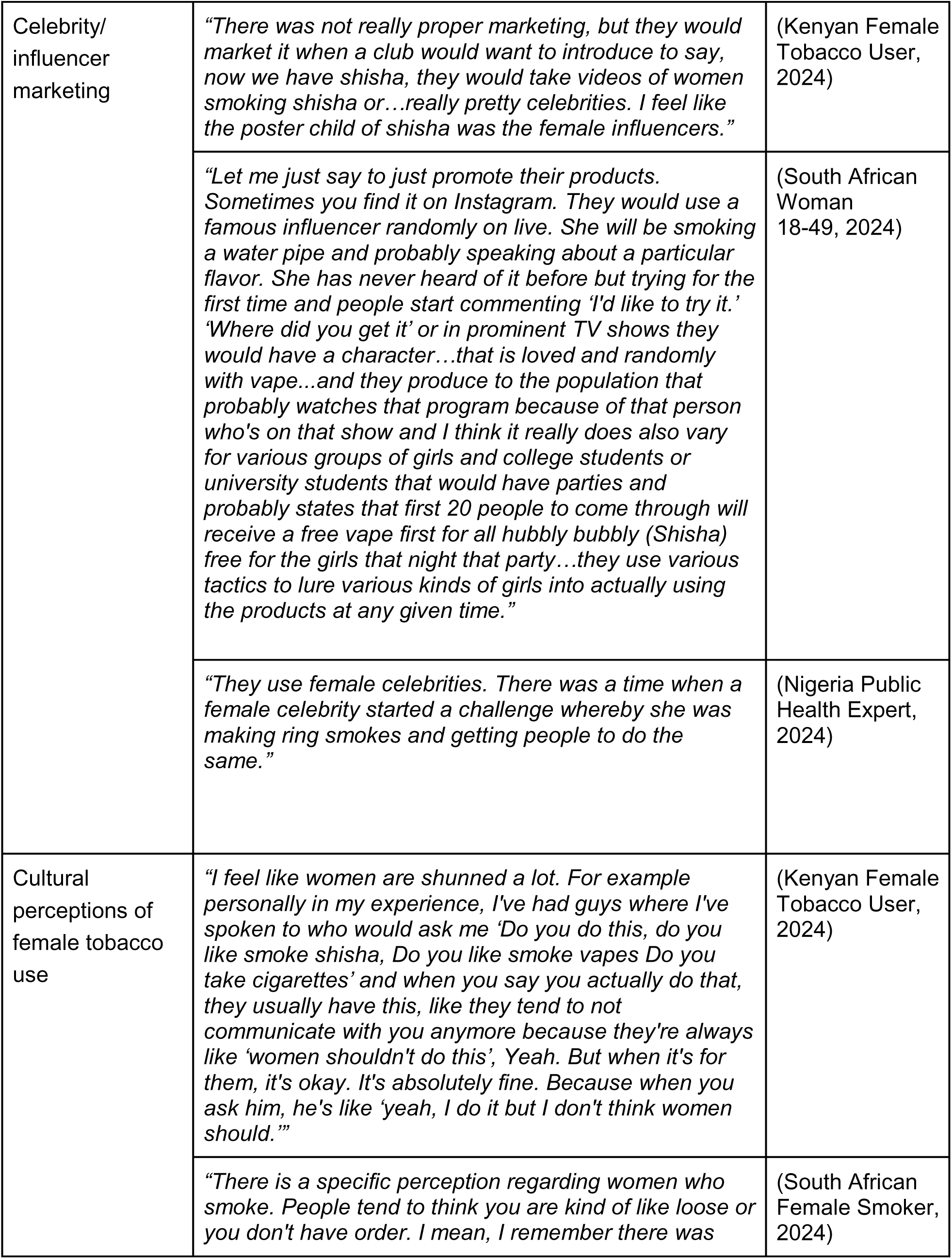
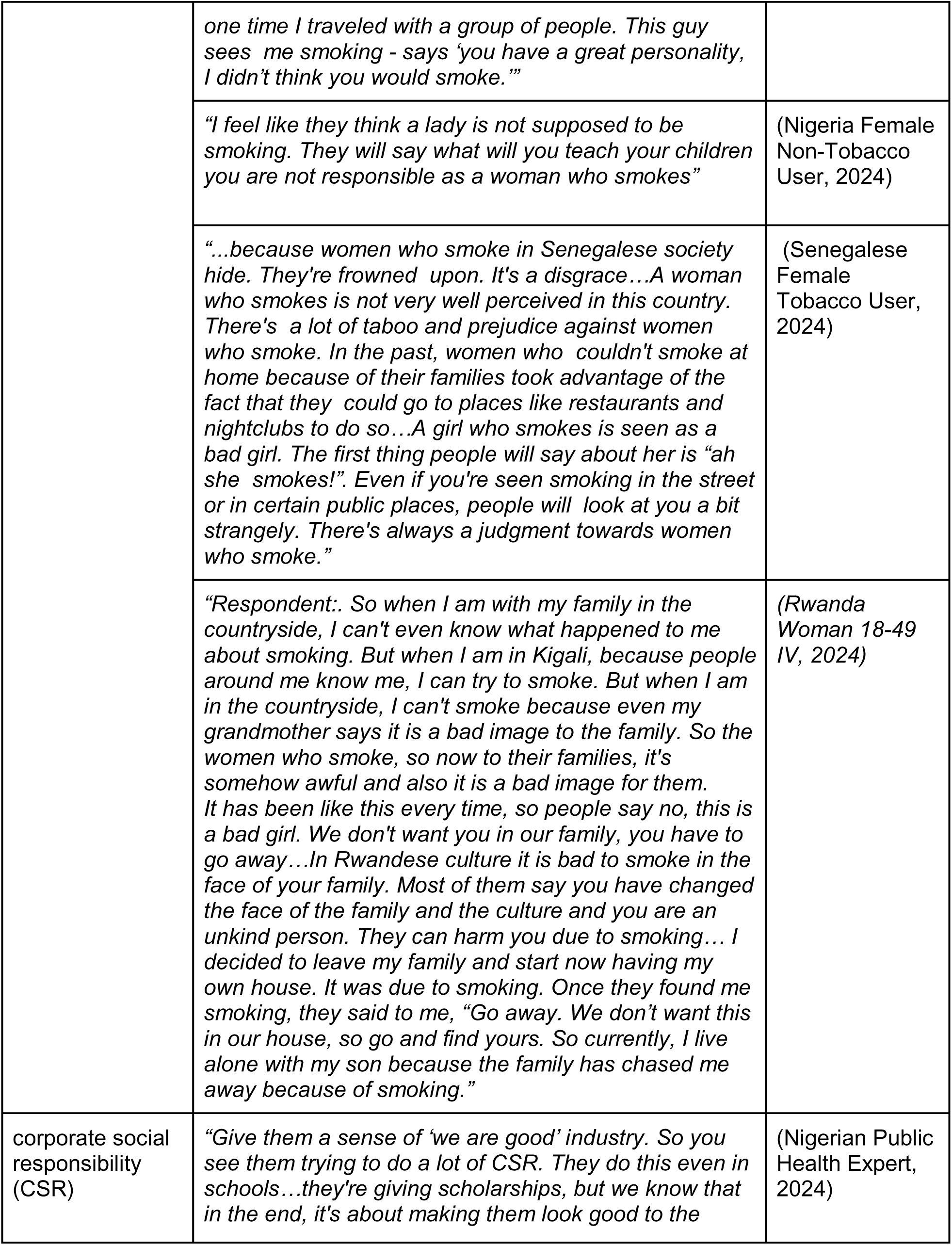
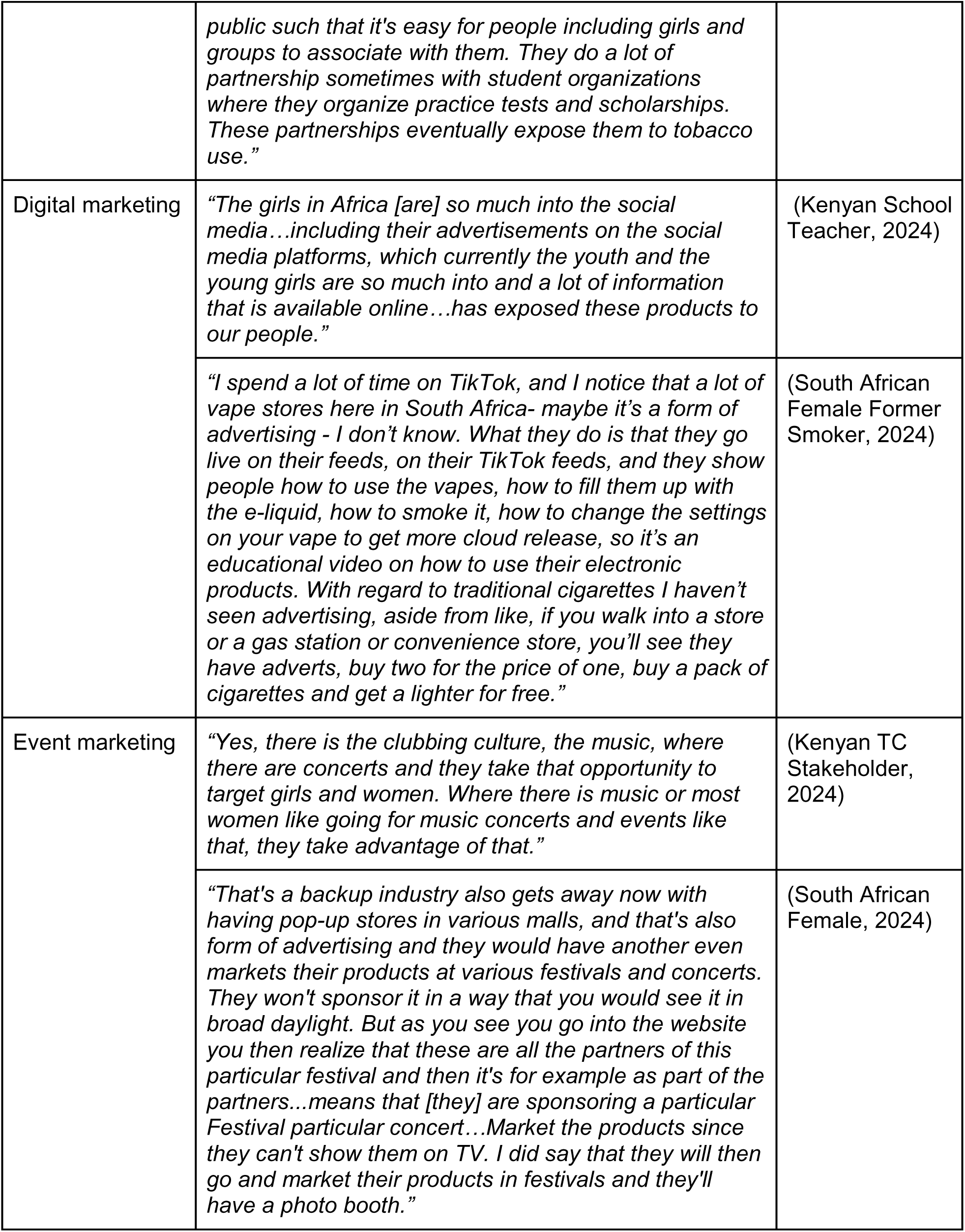
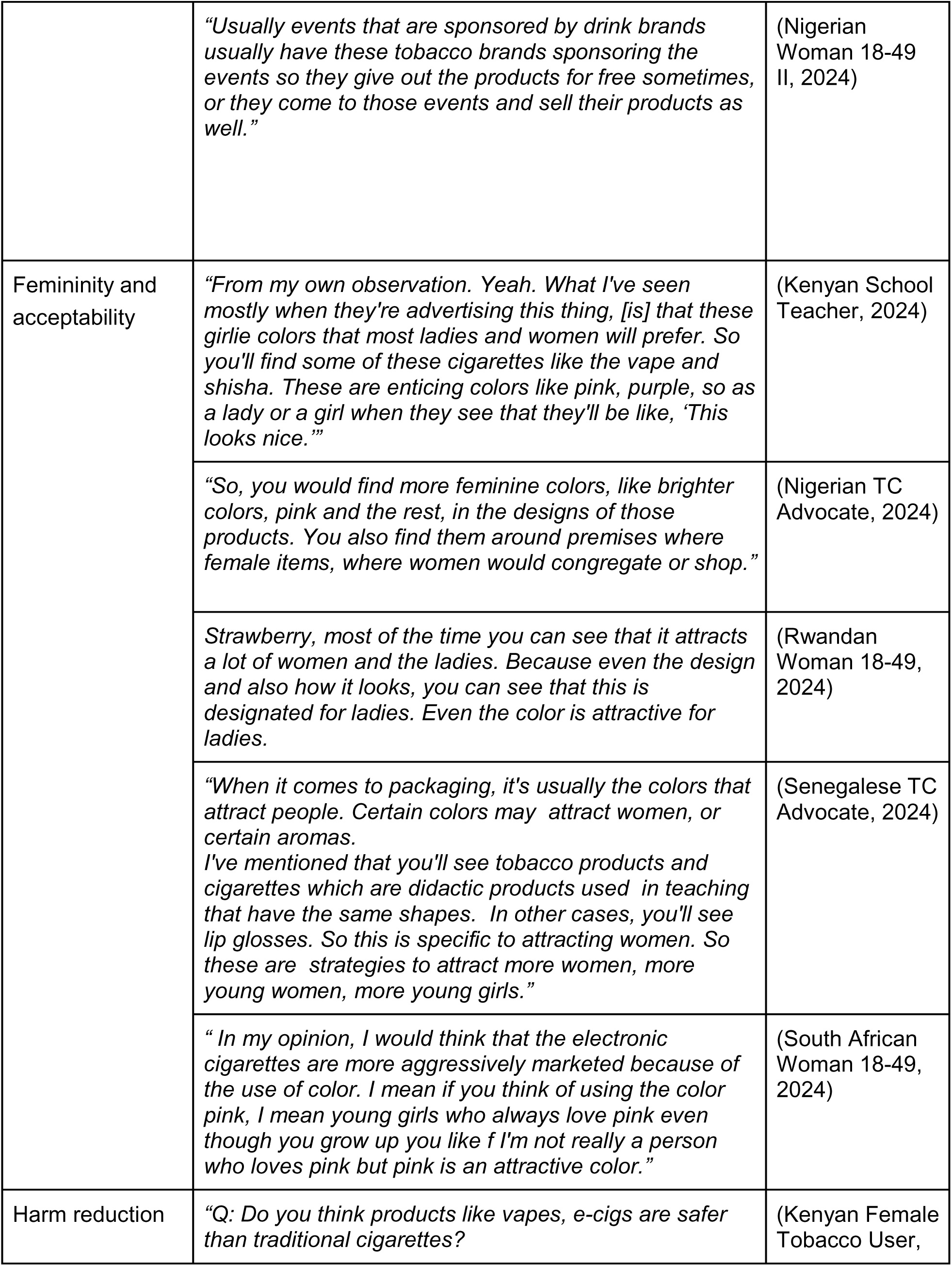
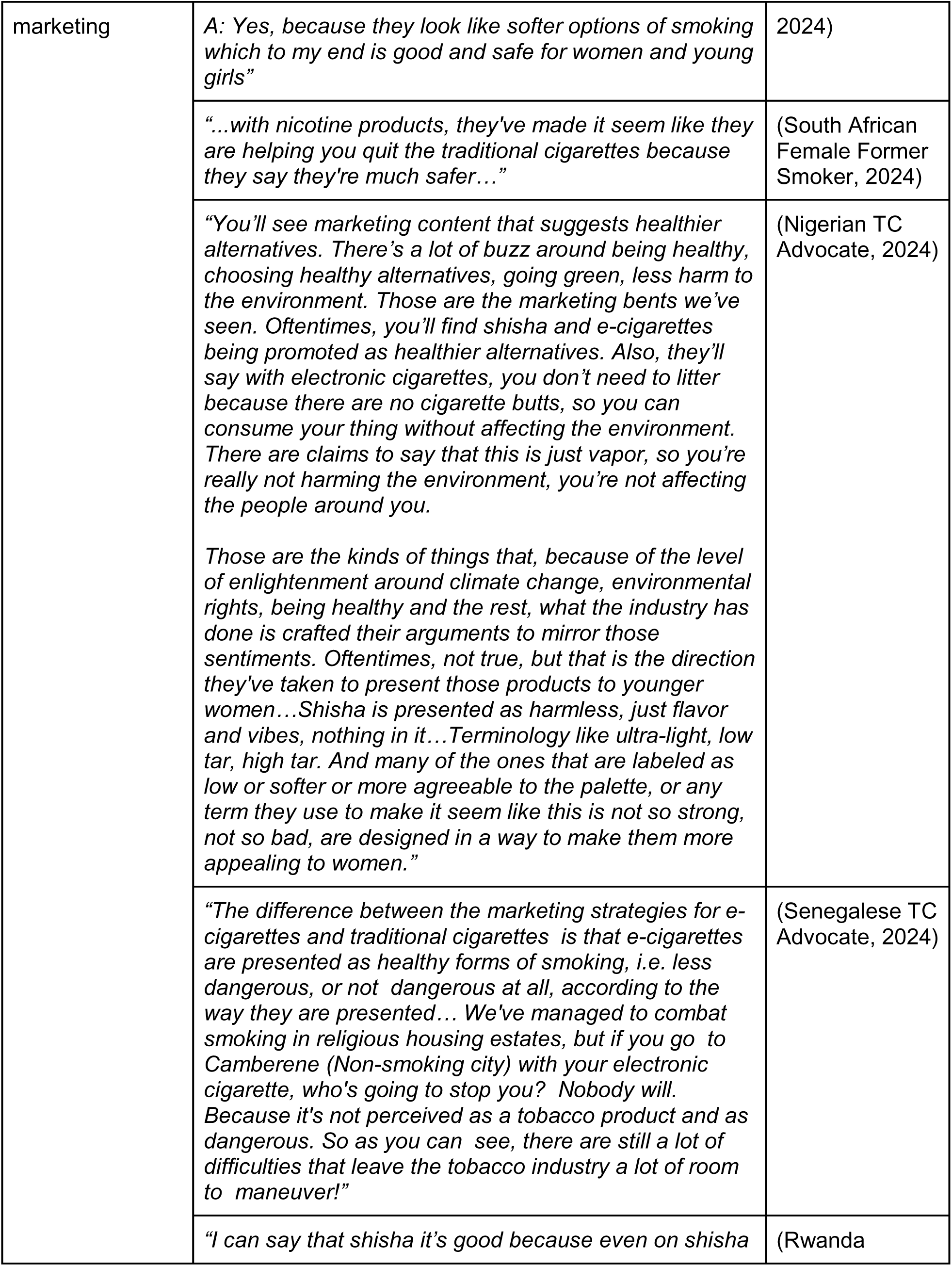
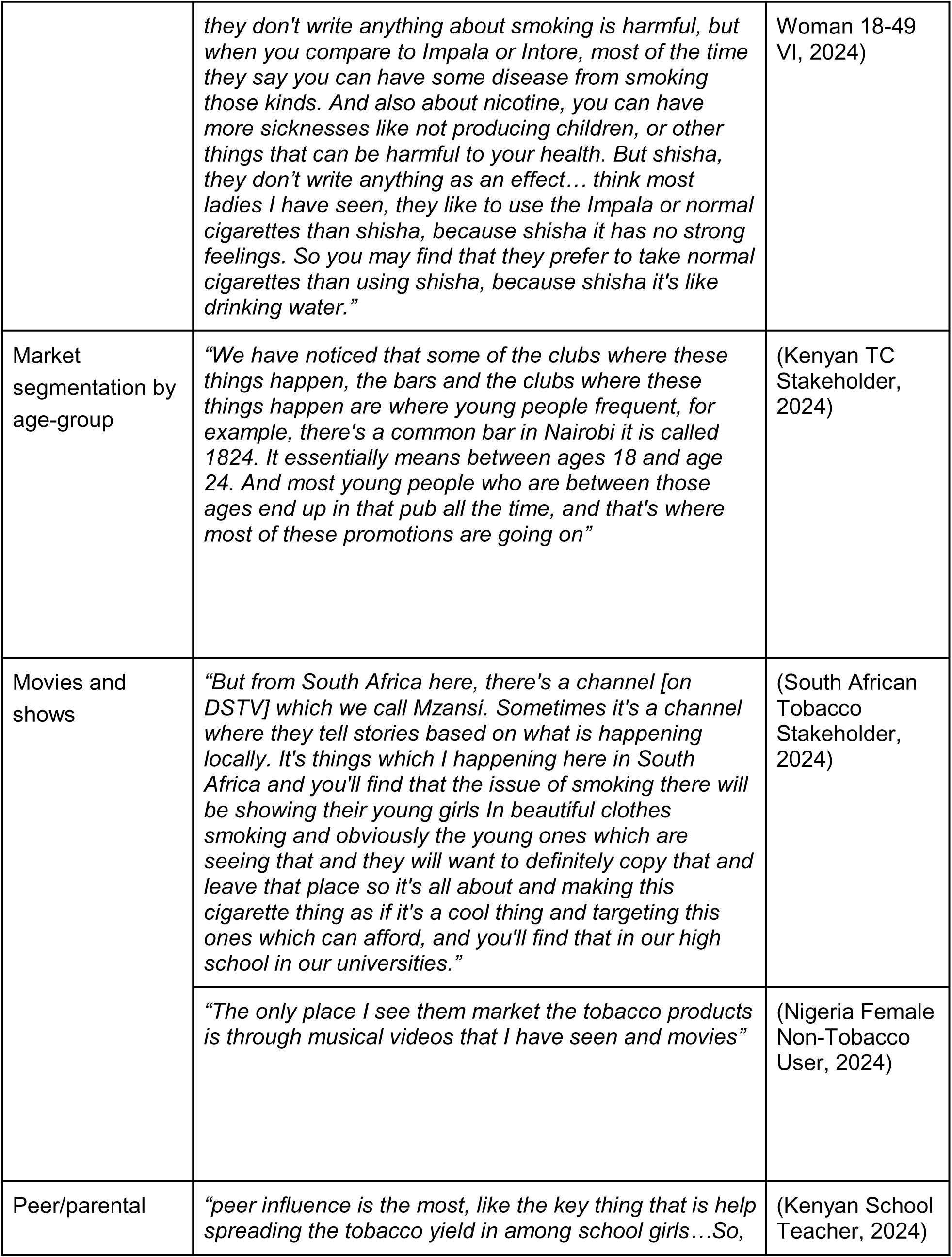
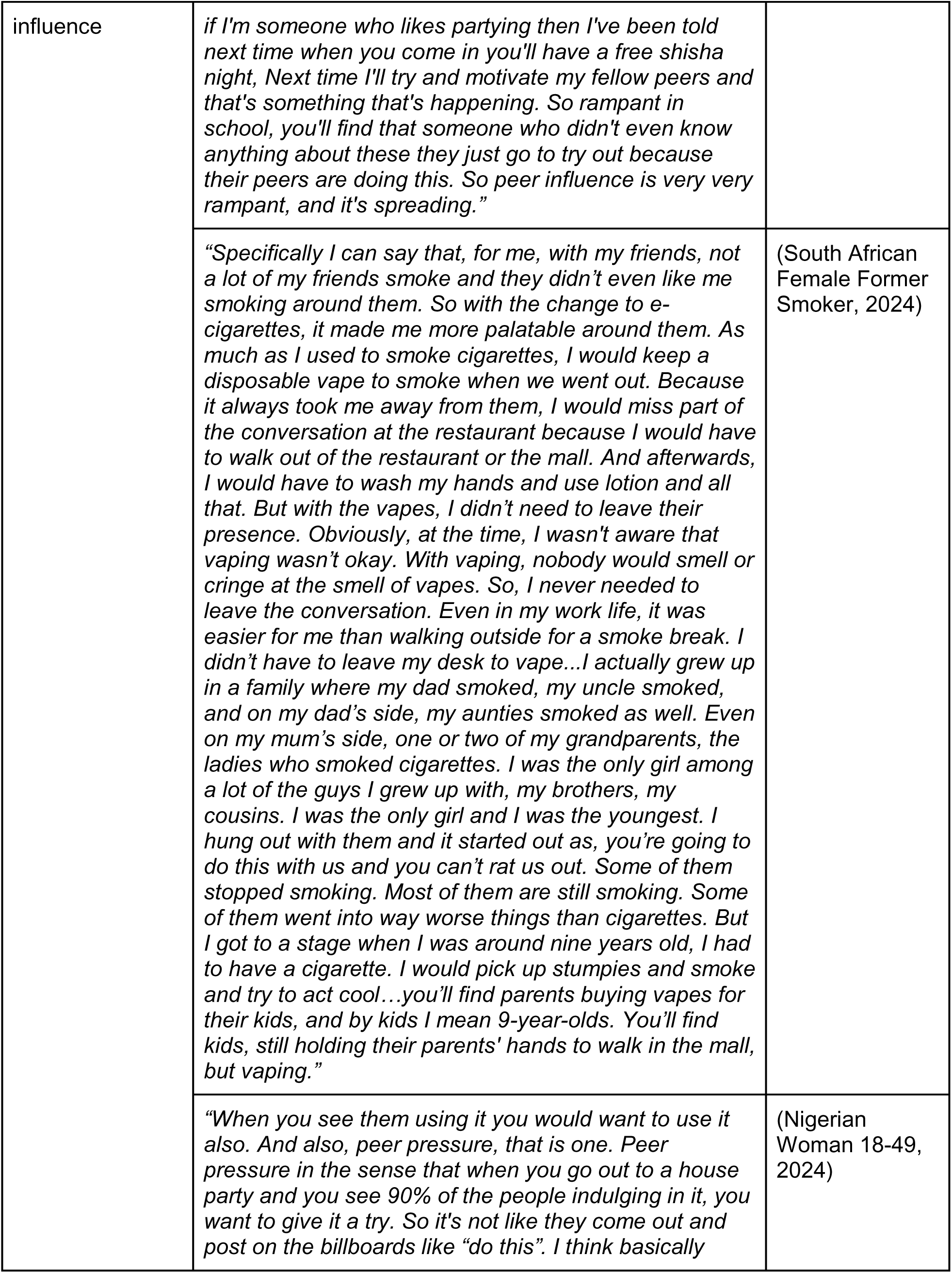
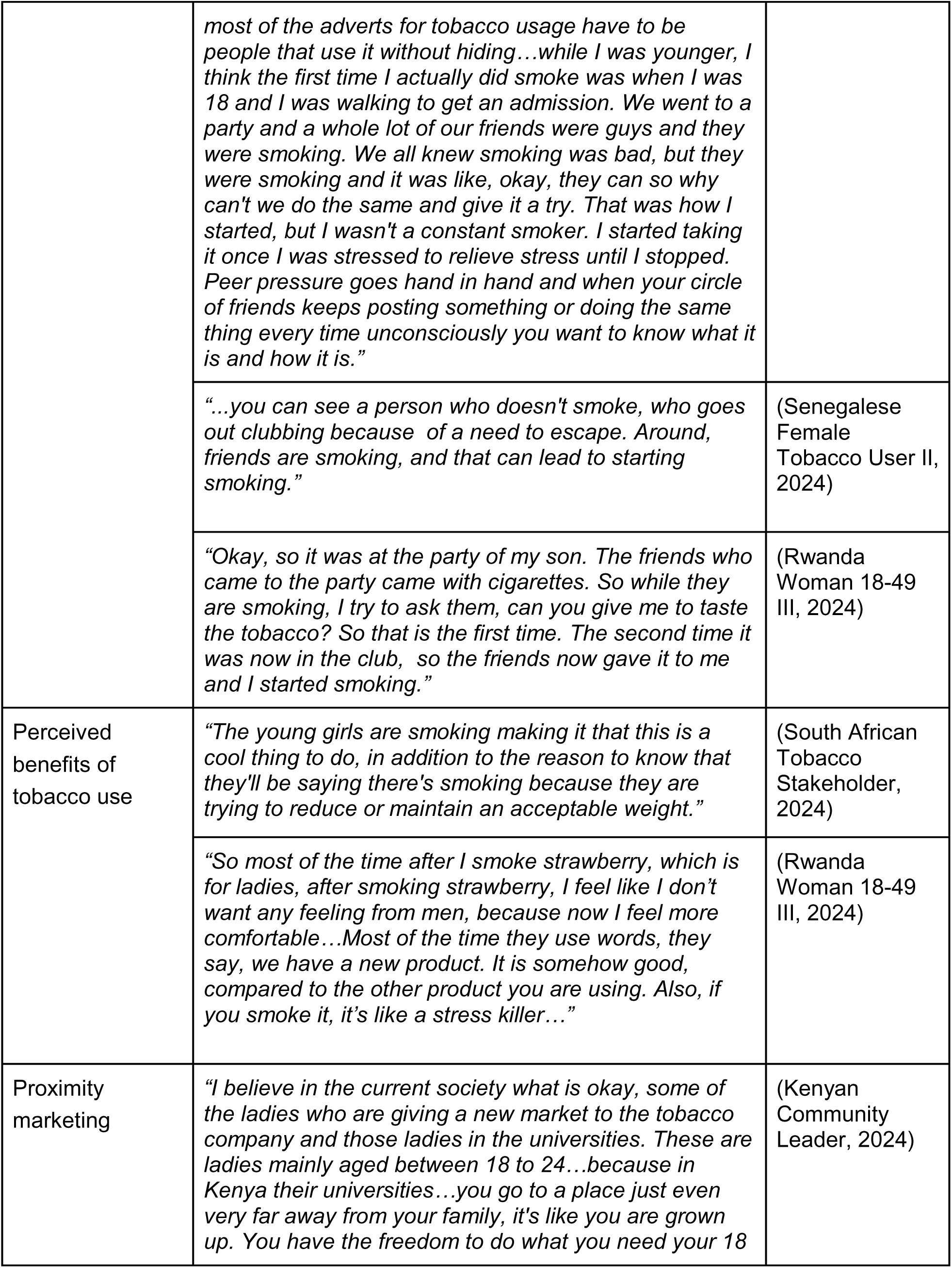
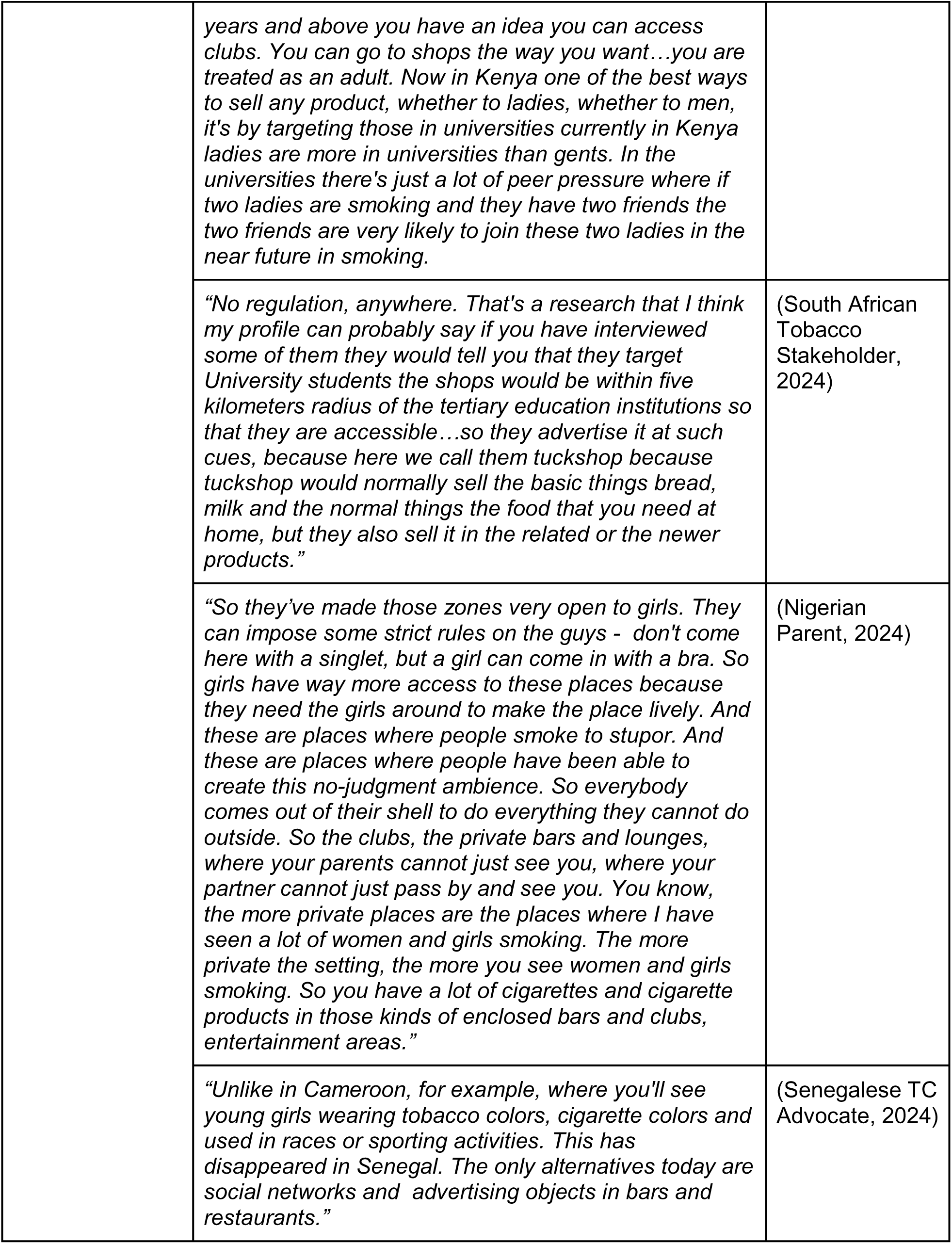
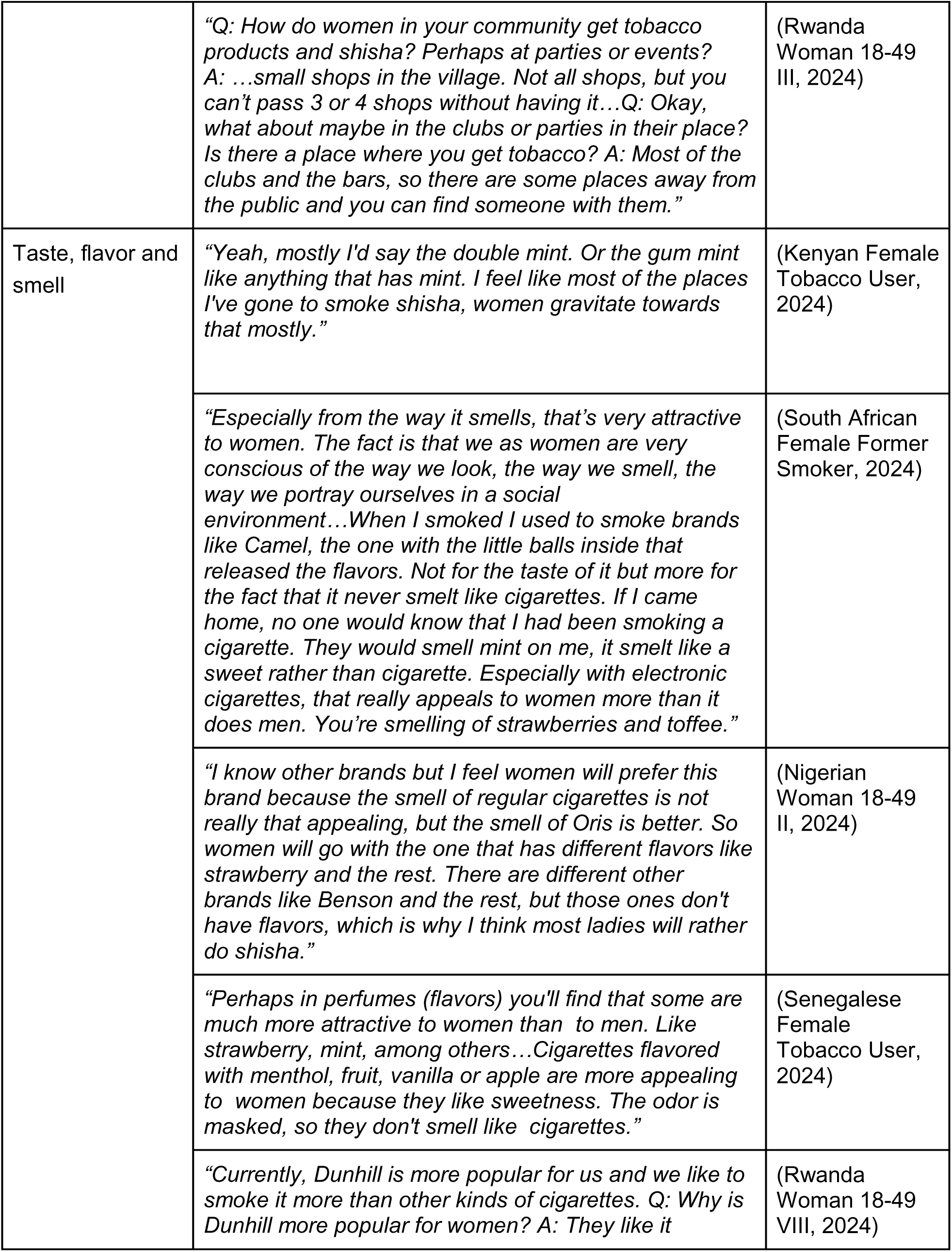
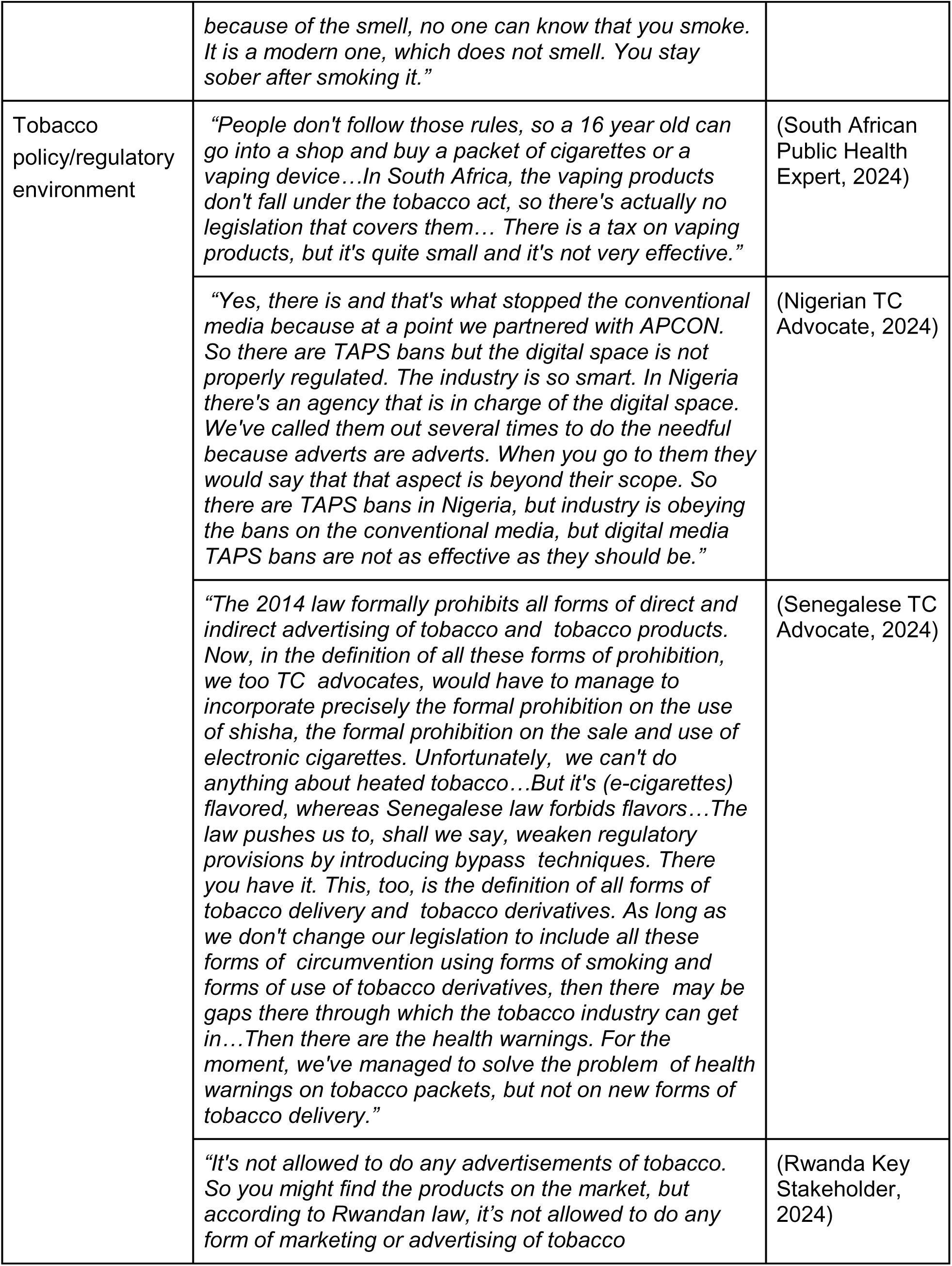
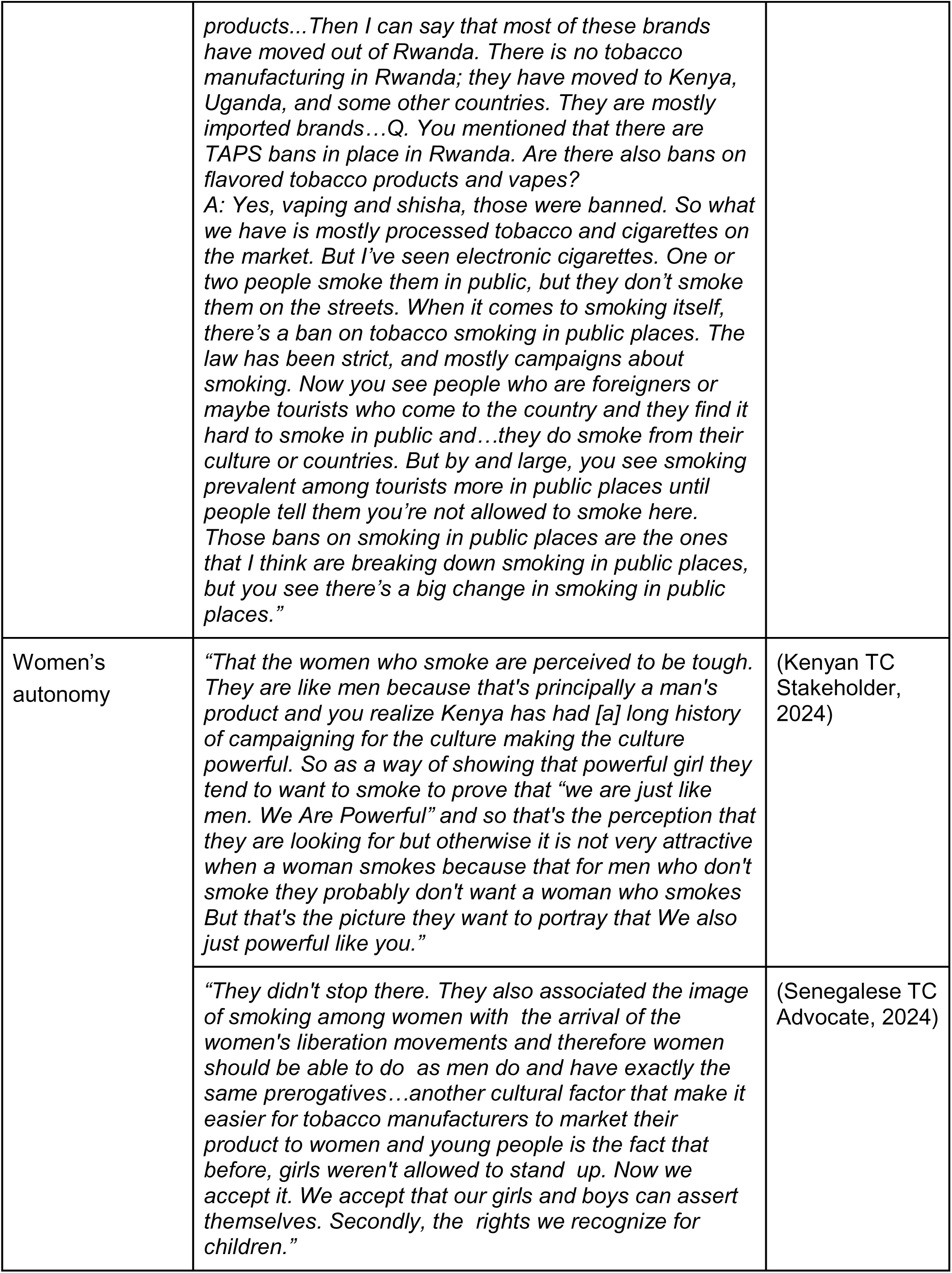
Results of Qualitative Analysis of KIIs.

